# Engineering Highly Thermostable Cas12b via De Novo Structural Analyses for One-Pot Detection of Nucleic Acids

**DOI:** 10.1101/2022.10.02.22280626

**Authors:** Long T. Nguyen, Santosh R. Rananaware, Lilia G. Yang, Nicolas C. Macaluso, Julio E. Ocana-Ortiz, Katelyn S. Meister, Brianna L.M. Pizzano, Luke Samuel W. Sandoval, Raymond C. Hautamaki, Zoe R. Fang, Sara M. Joseph, Grace M. Shoemaker, Dylan R. Carman, Liwei Chang, Noah R. Rakestraw, Jon F. Zachary, Sebastian Guerra, Alberto Perez, Piyush K. Jain

## Abstract

Clustered Regularly Interspaced Short Palindromic Repeats (CRISPR)-based diagnostics have elevated nucleic acid detection in terms of sensitivity, specificity, and rapidity in recent years. CRISPR-Cas systems can be combined with a pre-amplification step in a one-pot reaction to simplify workflow and reduce carryover contamination. Here, we report an engineered Cas12b system from *Brevibacillus* (eBrCas12b) with improved thermostability that falls within the optimal range (60-65°C) of the Reverse Transcription-Loop-Mediated Isothermal Amplification (RT-LAMP). Using *de novo* structural analyses via DeepDDG and HotSpot Wizard based on Alpha Fold and SWISS-MODEL predicted structures, mutations were introduced into the REC and RuvC domains of wild-type BrCas12b to tighten the hydrophobic cores of the protein, thereby enhancing its stability at high temperatures. We expressed, purified, and systematically characterized 49 BrCas12b variants with an emphasis on functionality and thermostability. The assay utilizing eBrCas12b, which we coined SPLENDID (**<u>S</u>**ingle-**<u>p</u>**ot **<u>L</u>**AMP-mediated **<u>e</u>**ngineered BrCas12b for **<u>n</u>**ucleic acid **<u>d</u>**etection of **i**nfectious **<u>d</u>**iseases), exhibits robust *trans-cleavage* activity up to 67°C in a one-pot setting—4°C and 7°C higher than wild-type BrCas12b and AapCas12b, respectively. We further validated SPLENDID clinically in 40 Hepatitis C (HCV) positive and 40 negative serum samples. A specificity of 97.5%, an accuracy of 90.0%, and a sensitivity of 82.5% were achieved. Results can be obtained via one-pot testing in as little as 20 minutes. With the extraction process, the entire assay can be performed in under an hour. Therefore, we believe that SPLENDID has the potential to become a widely universal platform for the detection of infectious diseases.

## Introduction

Since the beginning of the COVID-19 pandemic, CRISPR-based diagnostic platforms have become a prominent detection technology that could identify the emergence of future pathogens via massive surveillance testing and potentially replace the traditional time-consuming Reverse Transcription - quantitative Polymerase Chain Reaction (RT-qPCR) method^1-5^. Recent advancements in CRISPR-based tests allow for the combination of a preamplification step, such as reverse transcription loop-mediated isothermal amplification (RT-LAMP), and a CRISPR reaction into a convenient one-pot detection assay^6^. RT-LAMP is a highly sensitive reaction that can result in detectable signals in as fast as 5-10 minutes^7^. However, since the optimal temperature for the RT-LAMP reaction is around 60°C-65°C^7-9^, coupling it with CRISPR-Cas systems has been less effective due to the lack of Cas thermostability in this range. Recently reported STOPCovid technology employed a thermostable Cas12b from *Alicyclobacillus acidiphilus* (AapCas12b), but one drawback of the system is that the Cas effector ceases to display enzymatic activity at temperatures higher than 60°C^6^. In our previous study, we implemented a thermostable Cas12b, which is derived from *Brevibacillus sp*. SYP-B805 (BrCas12b) and exhibited high enzymatic activity up to 63.4°C within a single-pot CRISPR-diagnostic assay for the detection of nucleic acids. Our one-pot assay successfully distinguished multiple SARS-CoV-2 Variants of Concerns (VOCs) with high sensitivity and specificity within 20 minutes^10^. However, while BrCas12b allowed for detection at a higher temperature than AapCas12b, the range of detection is still narrow. RT-LAMP primers are substantially dependent on temperature due to their loop-forming nature. Consequently, single-degree changes often result in significant improvement in target amplification, sensitivity, and specificity^7, 11^.

To allow for enzymatic activity at high temperatures such that complete coverage of the optimal range of RT-LAMP could be attained, we employed *de novo* structural analyses to engineering a more thermostable BrCas12b for better flexibility within the use of the one-pot assay. We employed the capabilities of AlphaFold and SWISS-MODEL to obtain predicted structures of wild-type BrCas12b for the visualization of amino acid interaction^12-17^. These structures were inputted into stability prediction tools HotSpotWizard and DeepDDG, which helped us gain insight into potential mutations that encourage thermostability^18, 19^. These tools have been particularly useful for the determination of hotspots and predicted ΔΔG to aid in protein engineering for enhanced stability, catalytic activity, and specificity. For instance, an engineered version of xylanase from *Gloeophyllum trabeum* was demonstrated to have an improved temperature optimum up to 80°C and enhanced stability at low pH (2 – 7.5) compared to the wild-type enzyme enabling applications in feed and brewing industries^20^. In another study, mutations D52N and S129A were introduced into pepsin based on ΔΔG calculations and the half-life of these mutants was increased significantly^21^. However, so far there have been no reports on improving the thermostability of Cas effectors for diagnostic purposes.

Here, we report engineered BrCas12b variants (eBrCas12b) that show up to a 13-fold increase in activity and melting temperatures of 10°C and 6°C higher than AapCas12b and wild-type BrCas12b, respectively. When combined in a one-pot reaction with RT-LAMP, the engineered BrCas12b with the highest thermostability was observed to have robust detection capability up to 67°C. Since RT-LAMP is typically performed between 60-65°C, the engineered BrCas12b variant enabled the combination of the RT-LAMP and CRISPR trans-cleavage assays to be carried out at a higher temperature range, allowing for more versatility and ease of primer design. The one-pot assay, which we coined SPLENDID (**<u>S</u>**ingle-**<u>p</u>**ot **<u>L</u>**AMP-mediated **<u>e</u>**ngineered BrCas12b for **<u>n</u>**ucleic acid **<u>d</u>**etection of **i**nfectious **<u>d</u>**iseases) can be performed in low-resource areas and offers robust sensitivity and specificity in as little as 20 minutes.

## Results

### Identification of stabilizing mutants of BrCas12b

Type V and VI CRISPR-Cas systems such as Cas12a, Cas12b, and Cas13a-d have been harnessed for the detection of nucleic acids^1, 22-25^. While Cas12a and Cas13a have been extensively deployed for on-site applications, Cas12b has recently emerged as a distinct class of enzyme, the majority of which are thermostable^26-28^. Unlike Cas12a, Cas12b requires both a crRNA and tracrRNA to carry out the cleavage of double-stranded DNA (dsDNA) and subsequent collateral cleavage of single-stranded DNA (ssDNA) (referred to as trans-cleavage activity) (Fig. 1a)^26, 28^. Although CRISPR-Cas systems can detect nucleic acids, they suffer from low sensitivity that lies within the picomolar range^23, 29^. Traditionally, CRISPR-based detection platforms are coupled with a pre-amplification step to increase the sensitivity of detection^1-3, 22^. Amplification-free methods have also been developed and successfully executed^30, 31^. However, they require multiple guide RNAs to have detectable signal readouts, and as a result, it can be challenging for these assays to distinguish subvariants for genotyping purposes.

**Figure 1.**
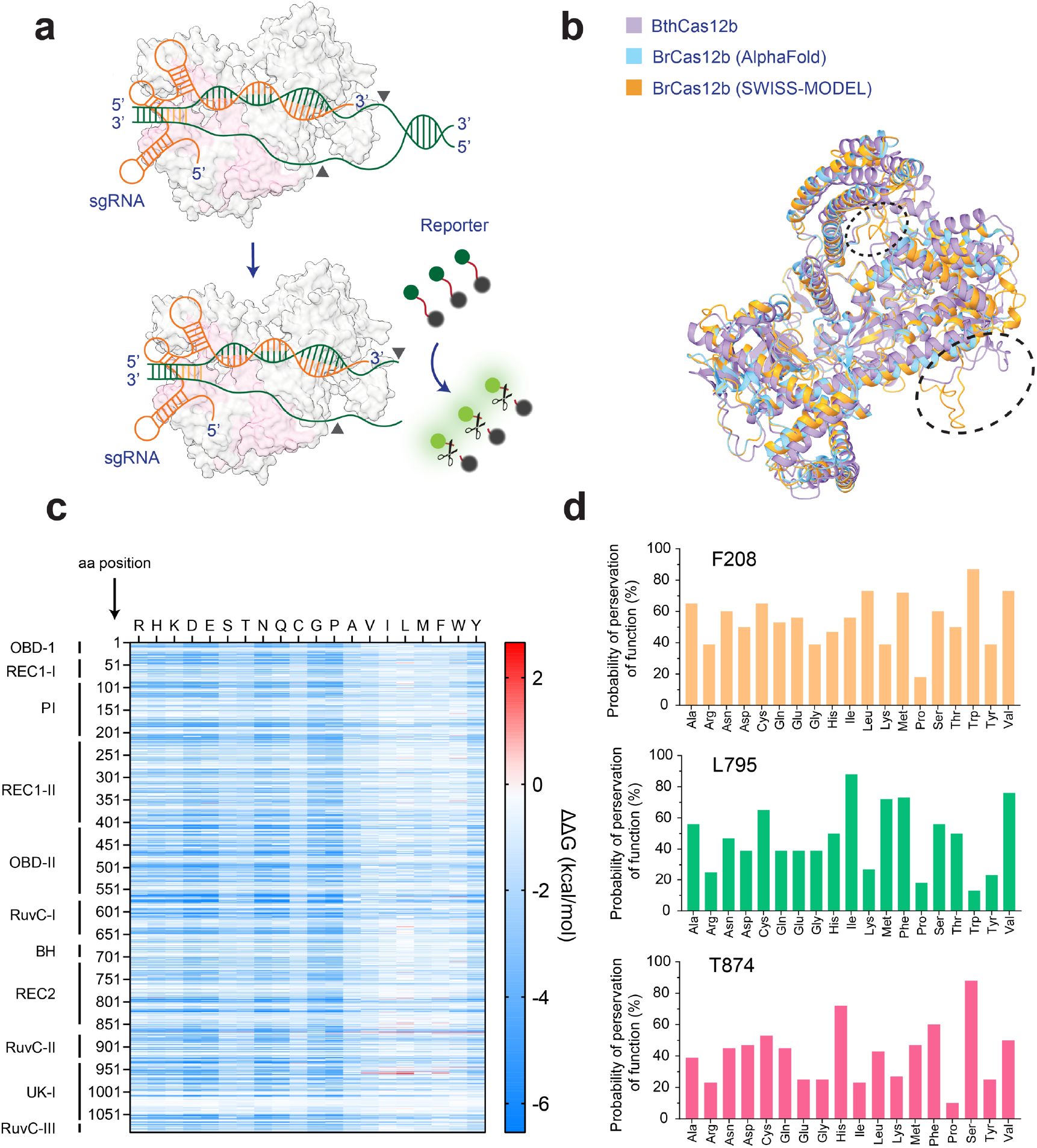
Structural prediction of BrCas12b. (**a**) Schematics of BrCas12b trans-cleavage. BrCas12b forms a binary complex with its sgRNA and binds to target dsDNA which activates its collateral ssDNA cleavage activity. (**b**) Predicted structures of BrCas12b derived from AlphaFold and SWISS-MODEL were spatially aligned with its most homologous Cas12b family member, BthCas12b (PDBID:5WTI)^38^. The circled regions indicate misalignment between the two predicted models when aligned to the BthCas12b structure. (**c**) The fitness landscape of BrCas12b was calculated by DeepDDG. Each tile represents a computationally predicted change in the free energy (ΔΔG) relative to the wild-type BrCas12b for the 20 amino acids. (**d**) Probability of preservation of function of three residues (F208, L795, and T874) which are likely to increase the thermostability of the BrCas12b effector calculated by Hotspot Wizard 3.0.

Reverse transcription - loop-mediated isothermal amplification (RT-LAMP) has emerged as a powerful tool for molecular diagnostics due to its rapid and isothermal amplification in as quick as 5-10 minutes^7^. Since four to six primers are needed to achieve such high sensitivity, false positives resulting from the primer-dimer formation are a major limitation of this technology^32, 33^. Recent efforts have coupled RT-LAMP with a thermostable CRISPR-Cas complex in a one-pot reaction to overcome this drawback^6^. The programmability of the CRISPR-Cas systems improves the specificity of detection. Upon amplification, correct base-pairing between the guide RNA and the amplified target serves as an additional checkpoint to provide accurate signal readouts. The one-pot reaction further eliminates carryover contamination that is frequently observed with two-pot RT-LAMP-coupled platforms^10^. Most RT-LAMP reactions work optimally between 60°C – 65°C^7, 11^; however, only a handful of Cas enzymes are functional at this temperature range^6, 34^. The discrepancy in reaction conditions and inhibitory effects between amplification and trans-cleavage has also been a major hurdle in developing such one-pot systems. Recently, we reported the use of a thermostable Cas12b from *Brevibacillus sp*. SYP-B805 (BrCas12b) that outperformed previous platforms in a one-pot detection setting. Nevertheless, the thermostable nature of BrCas12b only allowed this system to work robustly up to 63.4°C^10^.

Previous studies have shown that engineering the crRNA for Cas12a enhances its trans-cleavage activity^29, 35^. However, the thermostability of Cas enzymes is mainly linked to their secondary and tertiary structures^36, 37^. Therefore, rather than focusing on modifying sgRNA for BrCas12b, we aimed to engineer the enzyme itself not only to boost its activity but also its thermostability for better synergy with the RT-LAMP reaction. We employed structure-guided rational design to identify amino acid positions whose mutations are likely to be beneficial. To accomplish this, we predicted the structure of BrCas12b utilizing both AlphaFold and SWISS-MODEL followed by sequence alignment of these models with the most homologous solved Cas12b ortholog, BthCas12b (PDB ID: 5WTI)^12-15, 17, 38^, to identify functional domains. Overall, the AlphaFold and SWISS-MODEL predictions were congruent with one another except for two minor regions close to the RuvC domains (Fig. 1b). We next processed these predicted models via DeepDDG, a neural network-based algorithm to generate a fitness landscape of all amino acid positions^19^. We observed that the majority of the mutations were destabilizing except for hydrophobic substitutions in the REC1, RuvC, and UK-I regions (Fig. 1c). Based on Gibbs free energy changes from the fitness landscape, we analyzed these mutations individually on the AlphaFold structures to determine predictors of thermostability in terms of hydrophobicity, compactness, hydrogen bonding, and salt bridges. In addition to these thermostability factors, we also applied a semi-rational protein design tool called HotSpot Wizard 3.0 to select the best candidates for further validation^18^. We pooled a few single-point mutations that were predicted to be destabilizing according to their ΔΔG values as controls. We identified the three most promising candidates (F208, L795I, and T874) from a consensus of both algorithms and their computed mutational landscape, which inferred the probability of preservation of function (Fig. 1d).

### Engineered BrCas12b showed robust activity at high temperature

We expressed and purified 35 single-point mutated BrCas12b variants and characterized their thermostability and catalytic activity at different temperatures (Fig. S1). Using differential scanning fluorimetry, we observed that 16 variants increased the melting temperatures (T_m_) relative to the wild-type BrCas12b, whose apo form had a T_m_ of 61.4°C. Five variants (F208W, D567W, D868I, D868V, and D951F) displayed an increase of more than 2°C in their T_m_ compared to the wild type (Fig. 2a and Fig. S2). To investigate whether a combination of these stabilizing mutations would display an additive effect on the overall stability of the protein, we stacked beneficial mutations on top of each other to create several multi-mutated variants. We observed that the majority of stacked mutations such as F208W/D868V (FD), F208W/N524V/D868V (FND), F208W/N524V/T874H (FNT(H)), and F208W/N524V/L795I/D868V (FNLD) exhibited an overall additive increase of T_m_; however, there were a few exceptions—for instance, F208W/N524V/T874S (FNT(S)) and F208W/N524V/L795I/D868V/T874S/A1015E (FNLDTA) decreased thermostability upon stacking (Fig. 2a-c).

**Figure 2.**
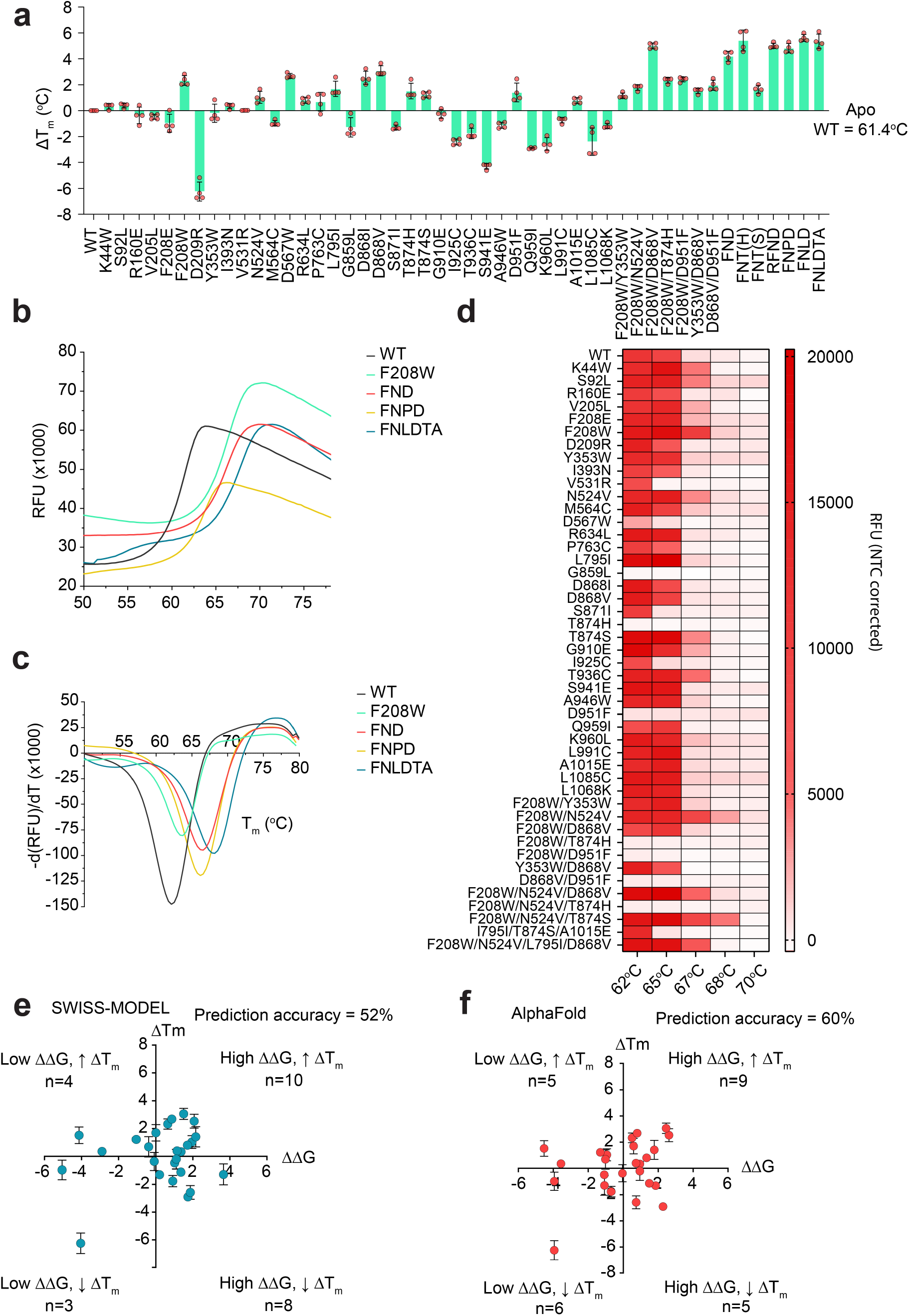
Functional characterization of BrCas12b variants. (**a**) Change in melting temperature of BrCas12b variants compared to the wild-type effector (n = 4, two technical replicates examined over two independent experiments). Error bars represent mean ± SEM. (**b**) Differential scanning fluorimetry plots in RFU comparing the wild-type BrCas12b for selected variants screened from (a) (n = 4, two technical replicates examined over two independent experiments). (**c**) Differential scanning fluorimetry plots in terms of derivative of RFU in (b). Global minima of the curve were determined to be the melting point of the protein (n = 4, two technical replicates examined over two independent experiments). (**d**) The trans-cleavage activity of BrCas12b variants was determined at increasing temperatures. Heat map represents the background corrected RFU at t = 15 min (n = 2 biological replicates). (**e**) and (**f**) Prediction accuracy in terms of thermostability for SWISS-MODEL and AlphaFold models (n=4, two technical replicates examined over two independent experiments).

We performed an *in vitro* trans-cleavage assay on all variants to check for functional preservation at elevated temperatures. While the wild-type BrCas12b variants ceased to work above 65°C (possibly due to denaturation at high temperature), the engineered BrCas12b variants FN, FND, FNT(S), FNLD, RFND, and FNLDTA showed robust activity up to 68°C (Fig. 2d and Fig. S3). We also observed that some stabilizing mutants like D567W, T874H, and D951F were no longer active; by examining the predicted structure, we learned that these mutations were part of the RuvC and UK-I regions and potentially had an important role in Cas12b-mediated cleavage activity. We next sought out to characterize the accuracy of both the AlphaFold and SWISS-MODEL predictions by comparing the computationally predicted single-point mutants with the experimental observations. For the SWISS-MODEL, only 13 out of 25 mutants were experimentally consistent with the software calculations, yielding a prediction accuracy of 52%. On the other hand, the AlphaFold model resulted in 15 out of 25 correct mutants, yielding a higher prediction accuracy of 62% (Fig. 2e,f and Fig. S4,S5).

### Engineered BrCas12b exhibited improved thermostability while maintaining specificity compared to its wild type

We next carried out time-dependent trans-cleavage assays to investigate the stability of BrCas12b variants compared to its wild-type. Ribonucleoprotein complexes (RNP) were incubated between 10 and 60 minutes at different temperatures ranging from 60°C to 75°C. The dsDNA target and fluorescence-based reporter were then added to the RNP complex to initiate non-specific ssDNA cleavage. Based on kinetic measurements, we observed that the FND and RFND (R160E/F208W/N524V/D868V) variants showed robust trans-cleavage activity at 68°C when the RNP complex was incubated for up to 20 minutes and 50 minutes, respectively. On the other hand, the wild-type BrCas12b failed to have detectable trans-cleavage activity under the same conditions (Fig. 3a-c). To investigate the specificity of these BrCas12b variants, we designed the dsDNA target with single-point or double-point mutations across its first 10 bases proximal to the PAM site. While the FND variants showed comparable specificity to the wild-type BrCas12b, the RFND displayed a much lower tolerance to double-point mutations near the seed region than observed in wild-type BrCas12b (Fig. 3d-f). Overall, the RFND variant exhibited the most thermostability and specificity among all the variants tested.

**Figure 3.**
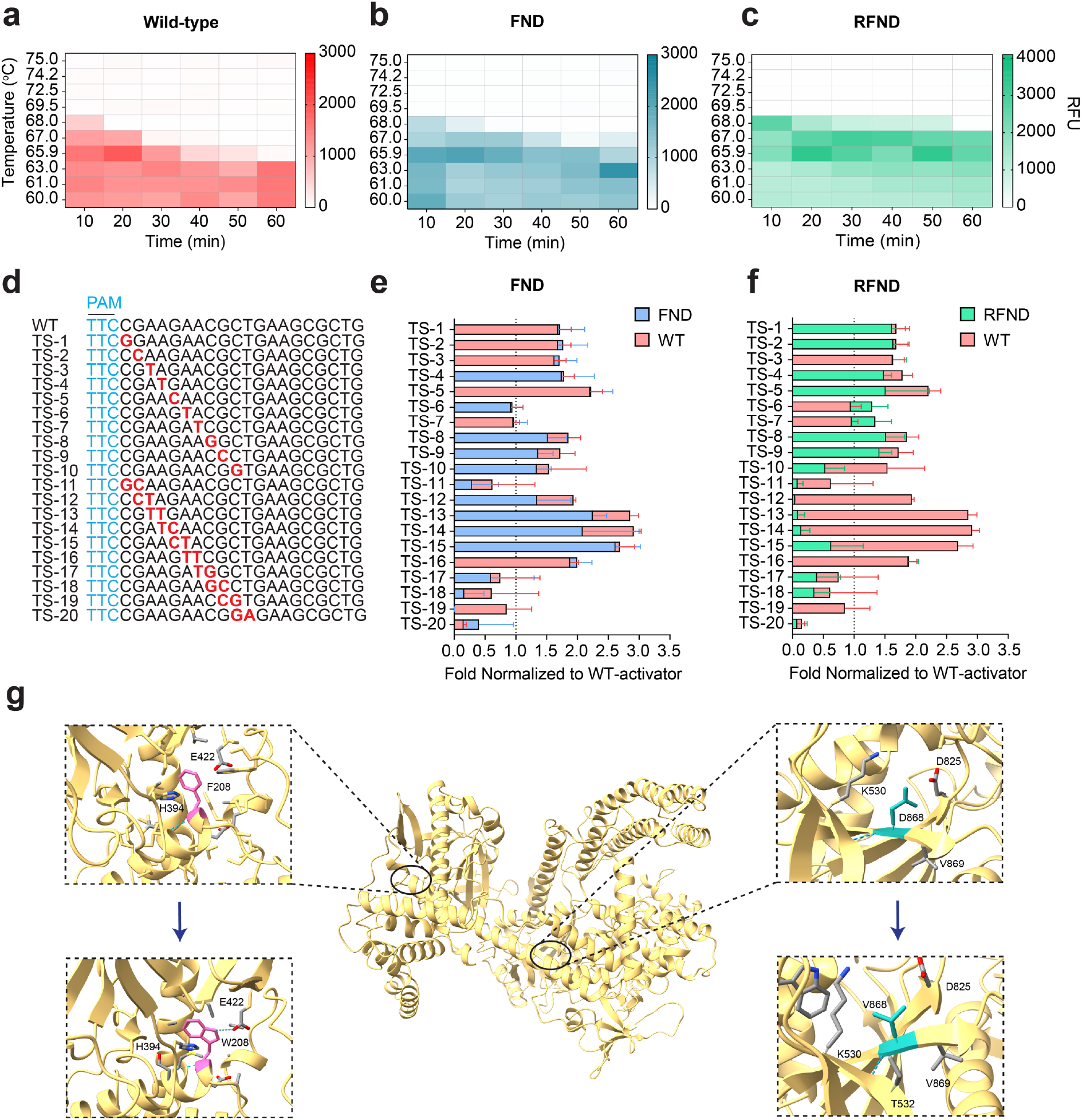
Engineered BrCas12b variants exhibit exceptional stability compared to wild-type BrCas12b. (**a**), (**b**), and (**c**) Time-dependent trans-cleavage activity at different temperatures for wild-type BrCas12b, FND (F208W/N524V/D868V) variant, and RFND (R160E/F208W/N524V/D868V) variant, respectively. The heat map represents background corrected RFU at t=15 min (n=2 biological replicates). (**d**),(**e**) and (**f**) Specificity testing of wild-type BrCas12b, FND variant, and RFND variant against mutated activators (n=2 biological replicates). (**g**) Structural insights into the mechanism of enhanced stability of BrCas12b variants at amino acid positions 208 and 868. The top panels display the wild-type BrCas12b with F208 (left) and D868 (right) while the bottom panels depict the engineered eBrCas12b with W208 (left) and V868 (right).

We hypothesized that the mutations F208W, N524V, and D868V contributed to the increase in melting temperature while the R160E mutation enhanced the enzyme activity. By looking at the BrCas12b predicted structure closely, we observed that the tryptophan at position 208 further stabilized the interaction with His394 likely by hydrophobic π–π stacking, and formed an extra hydrogen bond with Glu422^39^. Through biochemical assays, this mutation was observed to be the most contributing factor to the overall thermostability of the enzyme. Val868 allowed for improved compactness in the hydrophobic core of the RuvC region through interactions with more hydrophobic residues around it (Fig. 3g). For the mutation R160E, we did not observe an increase in the melting temperature via differential scanning fluorimetry; however, its trans-cleavage activity was higher than the wild-type BrCas12b. We hypothesized that the conversion of R160E to a negatively charged residue may have subsequently led to the enhancement in dsDNA cleavage.

### Engineered BrCas12b variants show robust activity up to 67°C in an RT-LAMP-mediated one-pot reaction

We aimed to investigate the thermostability of several engineered BrCas12b in a one-pot setting. To accomplish this, we employed a LAMP primer set described in Broughton et al.^2^ that was used to detect SARS-CoV-2 N-gene and tested them at high temperatures ranging from 62°C to 70°C with an increment of 1°C. Interestingly, these proof-of-principle primers worked robustly up to 68°C. We next coupled some of the most promising BrCas12b variants with the RT-LAMP reaction in a single pot at different temperatures ranging from 64°C to 68°C. Notably, FND, FNLD, RFND, FNPD, and FNLDTA variants showed high detection signal at 64°C with FND, FNLD, and RFND exhibiting an approximately 7-fold increase in activity compared to the wild-type BrCas12b (Fig. 4a). At 65°C, the wild-type BrCas12b completely ceased activity due to its denaturation at high temperatures (Fig. 4b). The engineered BrCas12b variants worked robustly up to 66°C but performed poorly at 67°C possibly due to reaching their thermostability limit (Fig. 4c,d).

**Figure 4.**
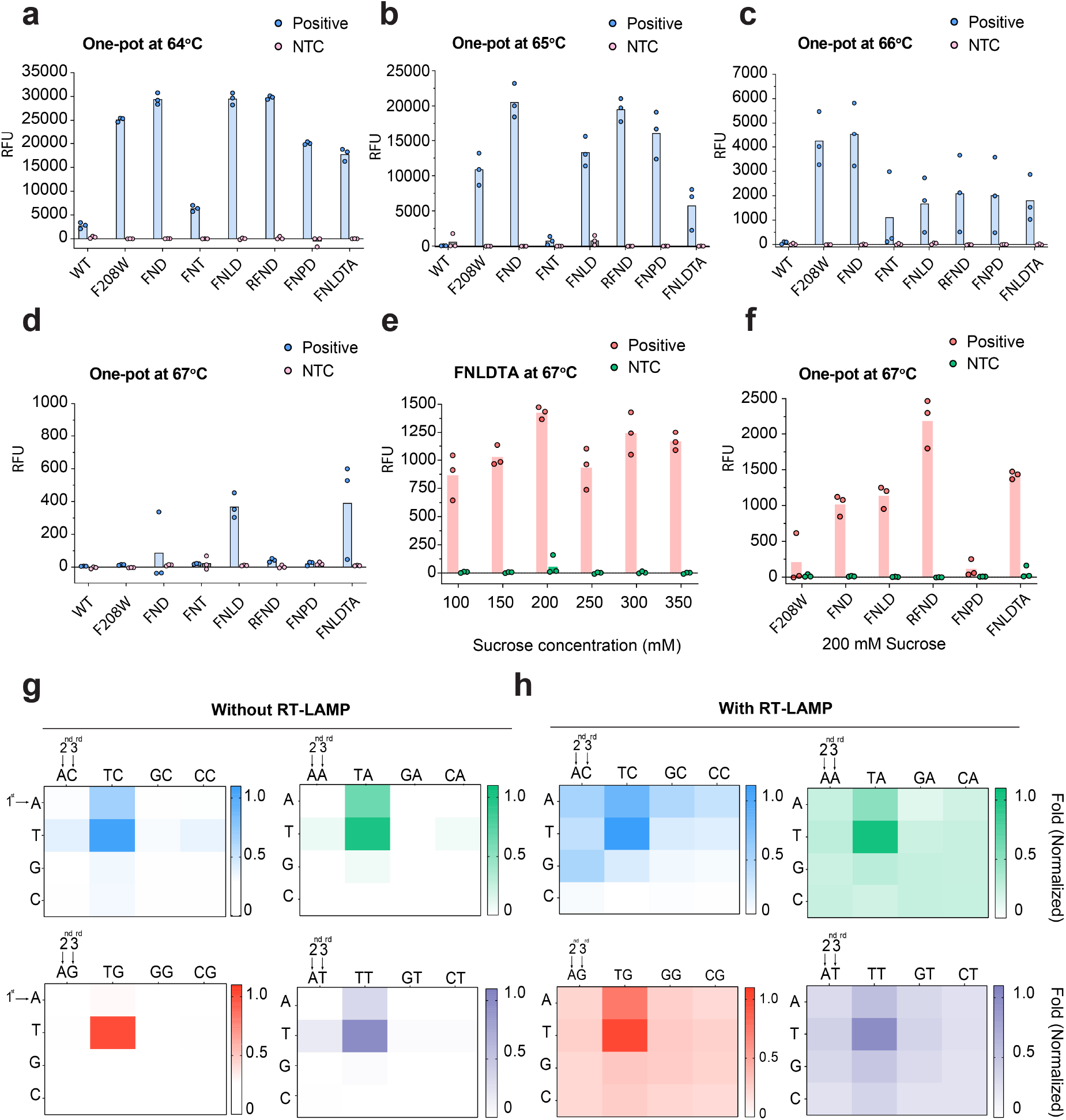
RT-LAMP-coupled one-pot detection of BrCas12b variants. (**a**), (**b**), (**c**), and (**d**) One-pot detection of BrCas12b variants at 64°C, 65°C, 66°C, and 67°C, respectively (n=3 biological replicates). The variants: FND, FNT, FNLD, RFND, FNPD, FNLDTA are abbreviated for F208W/N524V/D868V, F208W/N524V/T874S, F208W/N524V/L795I/D868V, R160E/F208W/N524V/D868V, F208W/N524V/P763C/D868V, F208W/N524V/L795I/D868V/T874S/A1015E, respectively. (**e**) Optimization of sucrose as an additive to the one-pot reaction at 67°C (n=3 biological replicates). (**f**) One-pot detection reaction of BrCas12b variants with 200 mM sucrose at 67°C (n=3 biological replicates). (**g**) and (**h**) PAM-dependent detection of RFND variant with and without RT-LAMP reaction.

Recent studies have shown that additives can boost the activity of CRISPR reaction in a one-pot setting^6, 40-43^. For instance, Joung et al. used taurine in their STOPCovid reaction and observed enhanced detectable signals^6^. Proline was also added to boost Cas12a activity in a two-pot setting^41^. Therefore, we sought to leverage the one-pot reaction using BrCas12b variants by exploring several additives such as taurine, mannitol, sucrose, trehalose, and betaine (Fig. S6). We selected the FNLDTA variant for testing as it showed the highest fluorescence signal at 67°C (Fig. 4d). We discovered that adding sucrose to the one-pot reaction at a final concentration of 200 mM greatly enhanced eBrCas12b enzymatic activity up to approximately 2.5-fold (Fig. 4e). We then applied 200 mM sucrose to the rest of the variants and found that sucrose was able to restore their activity at 67°C (Fig. 4f and Fig. S6). We coined the assay in which the RFND and optimized sucrose condition were used in a one-pot setting SPLENDID (**<u>S</u>**ingle-**<u>p</u>**ot **<u>L</u>**AMP-mediated **<u>e</u>**ngineered BrCas12b for **<u>n</u>**ucleic acid **<u>d</u>**etection of **i**nfectious **<u>d</u>**iseases).

CRISPR-Cas12b-mediated cleavage and detection of dsDNA are dependent on the presence of a short protospacer adjacent motif (PAM) upstream of the target sequence. It was empirically determined that the canonical PAM for BrCas12b was TTN^26-28, 44^. However, we speculated that when targeting single-stranded DNA (ssDNA), a PAM sequence may not be required as it shares similar enzymatic activity with Cas12a^23^. Since the RT-LAMP reaction can result in single-stranded DNA byproducts due to its loop-forming nature at elevated temperatures^7^, we hypothesized that PAMless or near-PAMless detection could be achieved by our RT-LAMP-coupled one-pot assay. This can potentially alleviate some challenges in primer and guide RNA designs. To investigate the PAM-dependency of eBrCas12b, we designed a PAM library of dsDNA activators that were comprised of all possible 3-nucleotide PAM sequences (NNN). We first sought to test the trans-cleavage of the RFND variant against these PAM combinations without the RT-LAMP step. We observed that the TTN-PAM-containing activators displayed the highest activity with the engineered Cas12b, consistent with previous studies. We also noticed significant trans-cleavage activity with activators containing the ATA, ATT, and ATC PAM but not with ATG. Similarly, TAA-, TAT-, and TAC-containing activators had low levels of detection, but not TAG. Our results indicate that the presence of a single T at either the first or the second position is sufficient to initiate the trans-cleavage of eBrCas12b provided that the third position contains A, T, or C. We next tested the detection of all the PAM-library activators in a one-pot setting (with RT-LAMP amplification). We observed that while the TTH PAM containing activators had the most robust detection, a majority of non-canonical PAM containing sequences also showed detection, albeit at lower levels—the only exceptions being CTC, CGC, and CCC PAMs, which showed no detection (Fig. 4g,h). Our results demonstrate that it is possible to target a variety of non-canonical PAM sequences if amplified by RT-LAMP in a one-pot assay.

### Validation of SPLENDID in Hepatitis C infected samples

Screening for Hepatitis C (HCV) is of importance for early treatment since there are no vaccines available^45-47^. Individuals infected with HCV can be cured within 8 to 12 weeks when detected early; otherwise, cirrhosis, hepatocellular carcinoma, and potential death can occur if left unnoticed^47, 48^. Traditional antibody assays offer up to 98% sensitivity and 99% specificity; however, these assays cannot distinguish between past infection and active infection. On the other hand, quantitative HCV RNA tests also exhibit similar sensitivity, specificity, and low limit of detection (10 – 15 IU/mL)^49-51^, but these detection platforms are laborious, time-consuming, and uneconomical. Therefore, as a proof of concept, we sought to develop a simple rapid test for Hepatitis C (HCV) using SPLENDID.

Prior to testing, a single-guide RNA was designed to target the 5’ untranslated region (5’ UTR) of the HCV RNA (+) genome (Fig. 5a). We then proceeded to clinically validate SPLENDID in 80 human serum samples (40 HCV-infected samples and 40 negative samples) with LAMP primers used in a study by Hongjaisee et. al^52^. Crude samples were processed via a magnetic-based extraction to isolate viral RNA. These samples were randomized and blinded before undergoing SPLENDID testing (Fig. 5b). All fluorescence-based measurements were compared to the RT-qPCR results to determine the sensitivity, specificity, and accuracy. Out of 80 samples, we observed 8 false negatives and 1 false positive, achieving a specificity of 97.5%, an accuracy of 90.0%, and a sensitivity of 82.5% (Fig. 5c,d). When analyzing the patient sample fluorescence results via a receiver operating characteristic curve (ROC) for the SPLENDID assay at t = 25 minutes, we observed that the area under the curve (AUC) was 92.3%, indicating a high threshold for differentiation between positive and negative samples (Fig. 5e).

**Figure 5.**
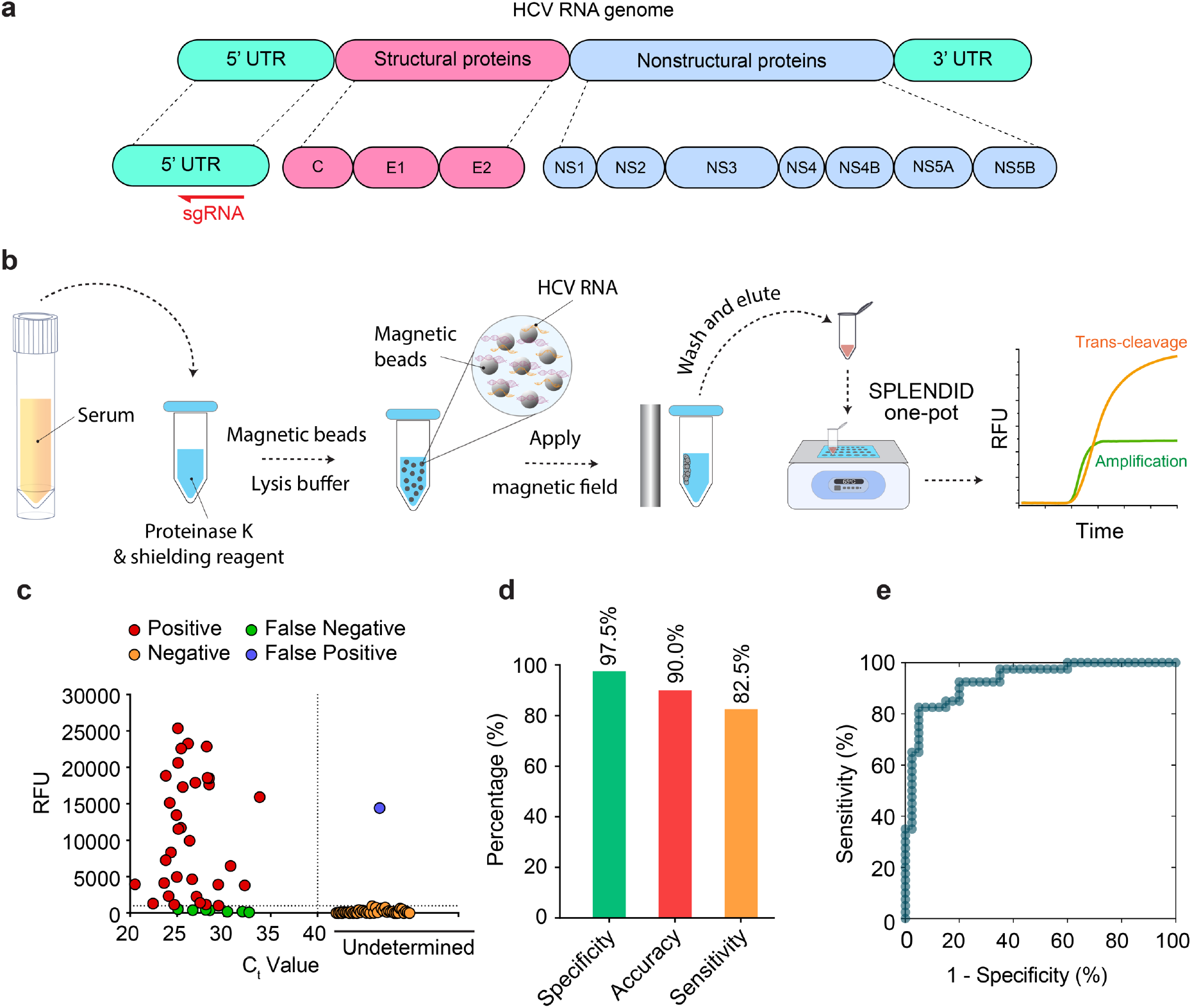
Clinical validation of SPLENDID in Hepatitis C infected samples. (**a**) Schematic of the Hepatitis C RNA genome. Single-guide RNA was designed to target the 5’ untranslated region (5’ UTR) of the virus. (**b**) Detailed steps of the SPLENDID assay from patient sample extraction via a magnetic-based method to the one-pot testing. (**c**) Fluorescence measurements of 80 HCV clinical samples from the SPLENDID assay. Signal readouts were taken at t = 25 minutes. (**d**) Summary of clinical validation SPLENDID in (d) in terms of sensitivity, specificity, and accuracy. (**e**) Receiver operating characteristic curve (ROC) of the assay at t = 25 minutes.

## Discussion

CRISPR-based diagnostics offers many advantages over traditional testing methods for nucleic acid detection due to its high specificity, flexible programmability, and quick turnaround time. To increase sensitivity, CRISPR reaction is often paired with a pre-amplification step such as RT-RPA or RT-LAMP. Here we report an engineered CRISPR-Cas12b system (eBrCas12b) with substantial improvements in terms of thermostability compared to other CRISPR-based one-pot assays.

Out of several mutations being introduced into the wild-type BrCas12b to create engineered variants, we observed a substantial improvement in thermostability of the proteins at two residues F208W and D868V, each with around a 2°C increase in melting temperature compared to the wild type. Structurally these two residues are located at two distinct regions of the protein, with F208 at the intersection of the PAM-interacting domain and REC1 and D868 being proximal to the catalytic pocket RuvCII domain. We hypothesized that F208W enabled π–π stacking with its neighbor H394 while D868V increased the compactness of its local region by increasing the hydrophobic surface interactions with its surrounding.

Our engineered detection assay using eBrCas12b, which we coined SPLENDID, works robustly up to 67°C in a one-pot setting, encompassing the optimal temperature range of the RT-LAMP reaction. SPLENDID combines two promising rapid detection platforms, amplification-based RT-LAMP and CRISPR assays, into one single pot reaction with improved synergy for a robust 2-step verification for the presence of target nucleic acids 1) amplification of the target and 2) collateral cleavage of BrCas12b. We applied SPLENDID to clinically validate 80 patient samples. Early detection of hepatitis C (HCV) is important for preventing more serious liver complications, such as liver cancer and cirrhosis. Current testing methods are costly, require specialized personnel and equipment, and may not distinguish between active and past infections. Our SPLENDID system utilizing our engineered thermostable BrCas12b (eBrCas12b) was able to successfully detect the presence of HCV in human serum samples with A specificity of 97.5%, an accuracy of 90.0%, and sensitivity of 82.5%. Compared to other available detection platforms for HCV, SPLENDID achieves a similar specificity; however, its sensitivity needs to be improved. We speculated that the sensitivity of our clinical validation testing heavily relied on the quality of the LAMP primer design. As a result, one way to improve the assay is to test many more LAMP primers set to obtain the most optimal one.

The SPLENDID system can be performed in as little as 20 minutes, and the whole process including sample extraction takes up to an hour. If paired with a more rapid extraction method, the overall assay duration can be reduced. Additionally, one-pot reagents can potentially be lyophilized as we discussed in our previous study, thus allowing for administration in low-resource areas^10^. We believe that the SPLENDID system has wide applications for the detection of emerging infectious diseases and can be quickly deployed to regions of interest at a low cost.

## Methods

### Computational Analyses

Protein structure predictions were performed on a local install of ColabFold with “AlphaFold2-ptm” parameters using A100 GPUs, where MSA search was done by MMseqs2 on the UniRef100 database^53^. For SWISS-MODEL predictions, protein sequences were input into the web-based tool via Expasy web server^13, 15-17, 54^. Protein structures in PDB files were processed via both HotSpot Wizard 3.0^18^ and DeepDDG^19^ web-based software.

### Nucleic Acid Preparation

Double-stranded DNA targets, single-guide RNAs, and fluorescence-quencher reporters were synthesized by Integrated DNA Technologies (IDT). PAM Library targets for the SARS-CoV-2 N gene were designed and synthesized by Twist Bioscience. For the trans-cleavage assay without RT-LAMP, the target-strand (TS) and non-target-strand (NTS) activators were ordered as 60-mer ssDNA oligos and annealed in a 5:1 (NTS:TS) ratio in 1X nuclease-free duplex buffer (IDT).

### Site-directed Mutagenesis

The wild-type BrCas12b gene was codon-optimized for *E. coli* expression, synthesized by Twist Bioscience, and cloned into a pET28a^+^ vector. For a mutant with single-point and double-point mutations, mutagenesis was carried out using the Q5® Site-Directed Mutagenesis Kit (New England Biolabs) following the manufacturer’s protocols. For simultaneous triple-point mutations, primers with intended mutations were designed and used to amplify each fragment of the BrCas12b using Q5® polymerase and assembled into a 6xHis-MBP destination vector (a gift from Scott Gardia, Addgene# 29656) using NEBuilder® HiFi DNA Assembly Kit (New England Biolabs).

### Protein Expression and Purification

Expression and purification of BrCas12b variants were performed according to a previous study with minor modifications^10^. After transforming BrCas12b variants into Rosetta™ 2(DE3)pLysS Singles Competent Cells (Millipore Sigma), individual colonies were selected and propagated overnight (12-15 hours) in 10 mL of Luria Broth (Fischer Scientific) containing the appropriate antibiotics. The bacterial cultures were then scaled up in a 2 L of Terrific Broth containing the corresponding antibiotics and 5 drops of antifoam 204 (Sigma Aldrich). Once an OD600 of 0.5-0.8 was attained, the culture was cooled in an ice bath for 30 mins, induced with 0.2 mM IPTG (isopropyl β-d-1-thiogalactopyranoside), and shaken at 18°C overnight (14-18 hours). The bacterial cells were then pelleted (39,800 xg at 4°C), diluted in lysis buffer (0.5 M NaCl, 50 mM Tris-HCl, pH = 7.5, 1 mM PMSF, 0.5 mM TCEP, 0.25 mg/ml lysozyme, and 10 μg/mL deoxyribose nuclease I), and disrupted by sonication. The lysed cells were then spun down (39,800 xg at 4°C), and the supernatant was subjected to suction filtration through a 0.22-micron filter (Millipore Sigma).

The filtered solution was purified using an FPLC Biologic Duoflow system (Bio-rad) through a Histrap 5 mL FF column (Cytiva), which was then washed (wash buffer: 0.5 M NaCl, 50 mM Tris-HCl, pH = 7.5, 20 mM Imidazole, 0.5 mM TCEP) before eluting (elution buffer: 0.5 M NaCl, 50 mM Tris-HCl pH = 7.5, 300 mM imidazole, 0.5 mM TCEP). For BrCas12b variants that were cloned into the 6xHis-MBP destination vector, an extra step of TEV site cleavage was performed to remove the MBP tag, and the protein was processed using Hitrap Heparin HP purification. The elution was evaluated using SDS-PAGE; the fractions containing the protein were consolidated and concentrated (2000 xg at 4°C) in a 50 kDa MWCO Amicon Ultra-15 centrifugal filter unit (Millipore Sigma). The protein was stored in -20°C for further use, with back-up reserves of each protein kept at -80°C after dilution (buffer C: 150 mM NaCl, 50 mM HEPES, pH = 7, 0.5 mM TCEP).

### Differential Scanning Fluorimetry

The melting temperature of each protein was mainly carried out in its apo form. BrCas12b was mixed in Protein Thermal Shift™ buffer (Thermo Fisher) in combination with 1x reaction buffer (100 nM NaCl, 50 nM Tris-HCl, pH=7.5, 1mM DTT, and 10 mM MgCl_2_) to a final concentration of 500 nM. Protein Thermal Shift dye™ (Thermo Fisher) was then added to each mixture before being transferred to the qPCR StepOne Plus system (Thermo Fisher). The temperature profile was observed over a temperature range of 25°C-80°C at a ramp rate of 1%/s. Duplicates of each experiment were performed in replicates of two.

### Temperature- and time-dependent trans-cleavage assay

The thermostable BrCas12b variants and sgRNA were combined in 1x NEBuffer 2.1 (New England Biolabs) to a final concentration of 50:100 nM (sgRNA:BrCas12b) and then transferred to a pre-heated CFX96 Real-Time PCR system with C1000 Thermal Cycler module (Bio-rad) using a temperature gradient setting across each row, ranging from 60°C to 75°C (60°C, 61°C, 63°C, 65.9°C, 69.5°C, 72.5°C, 74.2°C, and 75°C). The ribonucleoprotein complexes were incubated for 10 - 60 minutes with an increment of 10 minutes. A fluorescence-based reporter (FQ) and dsDNA activator were then added to each mixture to a final concentration of 250 nM and 1 nM, respectively. The reactions were isothermally incubated at their corresponding complexation temperature for consistency. Fluorescent measurements were taken every 30 seconds for 120 cycles on the HEX channel (λ_ex_: 525/10 nm, λ_em_: 570/10 nm). This experiment was repeated at incubation temperatures of 67°C and 68°C without the gradient set to cover a better temperature distribution. All experiments were repeated twice.

### One-pot RT-LAMP-coupled BrCas12b detection assay

LAMP primers were designed using NEB® LAMP Primer Design Tool from New England Biolabs (https://lamp.neb.com/#!/) and PrimerExplorer v5 from Eiken Chemical Co. (https://primerexplorer.jp/e/).

BrCas12b variants, single-guide RNA, fluorescence-quencher reporter, and 10x LAMP primers were combined in 1x Warmstart® Multi-Purpose/RT-LAMP Master Mix with UDG (New England Biolabs) to a final concentration of 200 nM, 400 nM, 2000 nM, and 1x LAMP primers (200 nM F3/B3, 1600 nM FIP/BIP, and 800 nM LF/LB), respectively, yielding a volume of 22 μL. The activator (dsDNA/RNA) was added to the mixture (25 μL final volume), and the reaction was then transferred to a CFX96 Real-Time PCR system with a C1000 Thermal Cycler module (Bio-rad). Fluorescence measurements were taken every 30 seconds per cycle for 120 cycles.

### PAM library

For trans-cleavage assays without the RT-LAMP step, BrCas12b and sgRNA activators were combined to a final concentration of 100 nM: 50 nM respectively in 1x NEBuffer 2·1 (New England Biolabs) and incubated at 65°C for 15 min. Fluorescence-based reporter (FQ) and 25 nM activators containing various PAM sequences were added to the reaction mixture containing the Cas12b: sgRNA complex to a final concentration of 250 nM and 1 nM respectively. The entire reaction was then immediately transferred to a Bio-rad CFX96 Real-Time system with a C1000 Thermal Cycler module. The reaction was isothermally incubated at 65 °C, and fluorescence measurements were read every 30 seconds per cycle over 120 cycles.

In the experiment with RT-LAMP, the one-pot BrCas12b detection assay method was utilized. 3 μL of 10 pM activators containing various PAM sequences were added to the reaction. Both experiments were carried out in duplicates and repeated twice.

### Patient samples extraction and processing

Patient sample collection and processing were approved by the University of Florida Institutional Review Board (IRB202003085). For clinical validation, 80 patient samples consisting of human serum were obtained, 40 of which were Hepatitis C positive and collected by HCV-TARGET; the other 40 deriving from healthy patients were collected by Boca Biolistics. Viral RNA was extracted from serum samples using Quick-DNA/RNA™ Viral MagBead Kit R2140 (Zymo Research. Extracted patient samples were chosen at random and blinded for one-pot testing.

The Quick-DNA/RNA™ Viral MagBead extraction was conducted according to manufacturer protocol. 10 μL of Proteinase K (20mg/mL) was combined with 200 μL of serum samples in 1.5 mL centrifuge tubes and incubated at room temperature (RT) for 15 minutes. DNA/RNA Shield™ (2x concentrate) was added to the serum sample containing Protease K in a 1:1 ratio. 800 μL Viral DNA/RNA Buffer was then incorporated into the combined mixture followed by the addition of 20 μL Magbinding Beads™ and vortexed for 10 minutes. The centrifuge tubes were placed in a magnetic stand until the beads were pelleted. The supernatant was aspirated and discarded. The beads were washed with 250 μL of MagBead DNA/RNA Wash 1, 250 μL MagBead DNA/RNA Wash 2, and two rounds of 250 μL of 100% ethanol. The tubes containing beads were air-dried for 10 minutes. DNA/RNA was eluted using 30 μL DNase/RNase-Free water and was subjected to a BrCas12b detection reaction.

### RT-qPCR of HCV clinical samples

HCV-specific primers and probe sequences were designed as described in Zauli et. al^55^. The probe was modified by adding an internal ZEN™ quencher from IDT to reduce the background signal. C_t_ values for 40 HCV infected and 40 healthy serum samples were obtained by using Luna® Probe One-Step RT-qPCR 4X Mix with UDG (NEB# M3019S) and following the manufacturer’s protocol. Briefly, Luna® Probe One-Step RT-qPCR 4X Mix with UDG, forward and reverse primers, and probe were combined to a final concentration of 1x, 0.4 μM (each), and 0.2 μM, respectively. Once assembled, the master mix was added to a 96-well plate (Applied Biosystems). 3 μL of the extracted patient sample were deposited in each well, to a total volume of 20 μL, to initiate the reaction. The 96-well plate was then inserted into the StepOnePlus Real-Time PCR system (Applied Biosystems), which set an auto threshold to calculate the Ct values.

## Data Availability

All data are presented in the main text as well as in the supplementary information. Predicted structures of all BrCas12b variants are included in the source files attached to this manuscript. All are welcome to contact the authors for additional data such as DeepDDG and Wizard Hotspot outputs.

## Acknowledgements

We are grateful to the entire HCV-TARGET consortium, particularly the principal investigator, Dr. David Nelson, and the staff member, Lauren Morelli, at the University of Florida for providing us with their expert guidance and clinical samples for this study. We are thankful to the past and current members of Jain Lab and Dr. Gary Wang’s Lab at the University of Florida for their insightful feedback. This work was financially supported by funds from the UF, the UF Herbert Wertheim College of Engineering, Dinesh O. Shah endowed professorship, NIH-NIAID R21AI156321, NIH-NIAID R21AI168795, and NIH-NIGMS R35GM147788. The funding sources did not have a role in the design of the study, the collection, analysis, or interpretation of data, nor in the writing of the manuscript.

## Author contributions

P.K.J., L.T.N., S.R.R., L.G.Y., and N.C.M. designed the experiments. L.T.N., S.R.R., L.G.Y., and N.C.M. mainly performed experiments with support from other co-authors. L.C. and A.P. performed AlphaFold predictions of all BrCas12b variants. L.T.N., S.R.R., and L.G.Y. wrote the initial manuscript with support from the rest of the co-authors. The manuscript was proofread, edited, and approved by all authors.

## References

1. Gootenberg, J.S. et al. Nucleic acid detection with CRISPR-Cas13a/C2c2. Science 356, 438-+ (2017).

2. Broughton, J.P. et al. CRISPR-Cas12-based detection of SARS-CoV-2. Nat Biotechnol 38, 870–874 (2020).

3. Patchsung, M. et al. Clinical validation of a Cas13-based assay for the detection of SARS-CoV-2 RNA. Nat Biomed Eng 4, 1140–1149 (2020).

4. Hampton, T. Virus Surveillance and Diagnosis With a CRISPR-Based Platform. Jama-J Am Med Assoc 324, 430–430 (2020).

5. Welch, N.L. et al. Multiplexed CRISPR-based microfluidic platform for clinical testing of respiratory viruses and identification of SARS-CoV-2 variants. Nat Med 28, 1083-+ (2022).

6. Joung, J. et al. Detection of SARS-CoV-2 with SHERLOCK One-Pot Testing. N Engl J Med 383, 1492–1494 (2020).

7. Notomi, T. et al. Loop-mediated isothermal amplification of DNA. Nucleic Acids Res 28, E63 (2000).

8. Amaral, C. et al. A molecular test based on RT-LAMP for rapid, sensitive and inexpensive colorimetric detection of SARS-CoV-2 in clinical samples. Sci Rep-Uk 11 (2021).

9. Huang, X., Tang, G.Y., Ismail, N. & Wang, X.W. Developing RT-LAMP assays for rapid diagnosis of SARS-CoV-2 in saliva. Ebiomedicine 75 (2022).

10. Nguyen, L.T. et al. A thermostable Cas12b from Brevibacillus leverages one-pot discrimination of SARS-CoV-2 variants of concern. Ebiomedicine 77 (2022).

11. Wong, Y.P., Othman, S., Lau, Y.L., Radu, S. & Chee, H.Y. Loop-mediated isothermal amplification (LAMP): a versatile technique for detection of micro-organisms. J Appl Microbiol 124, 626–643 (2018).

12. Jumper, J. et al. Highly accurate protein structure prediction with AlphaFold. Nature 596, 583-+ (2021).

13. Waterhouse, A. et al. SWISS-MODEL: homology modelling of protein structures and complexes. Nucleic Acids Research 46, W296–W303 (2018).

14. Kopp, J. & Schwede, T. The SWISS-MODEL repository: new features and functionalities. Nucleic Acids Research 34, D315–D318 (2006).

15. Guex, N., Peitsch, M.C. & Schwede, T. Automated comparative protein structure modeling with SWISS-MODEL and Swiss-PdbViewer: A historical perspective. Electrophoresis 30, S162–S173 (2009).

16. Studer, G. et al. QMEANDisCo-distance constraints applied on model quality estimation (vol 36, pg 1765, 2020). Bioinformatics 36, 2647–2647 (2020).

17. Bertoni, M., Kiefer, F., Biasini, M., Bordoli, L. & Schwede, T. Modeling protein quaternary structure of homo- and hetero-oligomers beyond binary interactions by homology. Sci Rep-Uk 7 (2017).

18. Sumbalova, L., Stourac, J., Martinek, T., Bednar, D. & Damborsky, J. HotSpot Wizard 3.0: web server for automated design of mutations and smart libraries based on sequence input information. Nucleic Acids Research 46, W356–W362 (2018).

19. Cao, H.L., Wang, J.X., He, L.P., Qi, Y.F. & Zhang, J.Z. DeepDDG: Predicting the Stability Change of Protein Point Mutations Using Neural Networks. J Chem Inf Model 59, 1508–1514 (2019).

20. Wang, X.Y. et al. A thermostable Gloeophyllum trabeum xylanase with potential for the brewing industry. Food Chem 199, 516–523 (2016).

21. Zhao, Y. et al. Rational Design of Pepsin for Enhanced Thermostability via Exploiting the Guide of Structural Weakness on Stability. Front Phys-Lausanne 9 (2021).

22. Gootenberg, J.S. et al. Multiplexed and portable nucleic acid detection platform with Cas13, Cas12a, and Csm6. Science 360, 439-+ (2018).

23. Chen, J.S. et al. CRISPR-Cas12a target binding unleashes indiscriminate single-stranded DNase activity. Science 360, 436-+ (2018).

24. Li, L.X. et al. HOLMESv2: A CRISPR-Cas12b-Assisted Platform for Nucleic Acid Detection and DNA Methylation Quantitation. ACS Synth Biol 8, 2228–2237 (2019).

25. Qiao, X. et al. Sensitive analysis of single nucleotide variation by Cas13d orthologs, EsCas13d and RspCas13d. Biotechnol Bioeng 118, 3037–3045 (2021).

26. Strecker, J. et al. Engineering of CRISPR-Cas12b for human genome editing. Nat Commun 10 (2019).

27. Tian, Y. et al. A novel thermal Cas12b from a hot spring bacterium with high target mismatch tolerance and robust DNA cleavage efficiency. Int J Biol Macromol 147, 376–384 (2020).

28. Teng, F. et al. Repurposing CRISPR-Cas12b for mammalian genome engineering. Cell Discov 4, 63 (2018).

29. Nguyen, L.T., Smith, B.M. & Jain, P.K. Enhancement of trans-cleavage activity of Cas12a with engineered crRNA enables amplified nucleic acid detection (vol 11, 4906, 2020). Nat Commun 11 (2020).

30. Fozouni, P. et al. Amplification-free detection of SARS-CoV-2 with CRISPR-Cas13a and mobile phone microscopy. Cell 184, 323-+ (2021).

31. Liu, T.Y. et al. Accelerated RNA detection using tandem CRISPR nucleases (vol 17, pg 982, 2021). Nat Chem Biol 17, 1210–1210 (2021).

32. Coelho, B.D. et al. Essential properties and pitfalls of colorimetric Reverse Transcription Loop-mediated Isothermal Amplification as a point-of-care test for SARS-CoV-2 diagnosis. Mol Med 27 (2021).

33. Hardinge, P. & Murray, J.A.H. Reduced False Positives and Improved Reporting of Loop-Mediated Isothermal Amplification using Quenched Fluorescent Primers. Sci Rep-Uk 9 (2019).

34. Ali, Z. et al. iSCAN: An RT-LAMP-coupled CRISPR-Cas12 module for rapid, sensitive detection of SARS-CoV-2. Virus Res 288 (2020).

35. Kim, H. et al. Highly specific chimeric DNA-RNA-guided genome editing with enhanced CRISPR-Cas12a system. Mol Ther-Nucl Acids 28, 353–362 (2022).

36. Hand, T.H., Das, A. & Li, H. Directed evolution studies of a thermophilic Type II-C Cas9. Method Enzymol 616, 265–288 (2019).

37. Kumar, S., Tsai, C.J. & Nussinov, R. Factors enhancing protein thermostability. Protein Eng 13, 179–191 (2000).

38. Wu, D., Guan, X.Y., Zhu, Y.W., Ren, K. & Huang, Z.W. Structural basis of stringent PAM recognition by CRISPR-C2c1 in complex with sgRNA. Cell Res 27, 705–708 (2017).

39. Fernandez-Recio, J., Vazquez, A., Civera, C., Sevilla, P. & Sancho, J. The tryptophan/histidine interaction in alpha-helices. J Mol Biol 267, 184–197 (1997).

40. Louwrier, A. & van der Valk, A. Can sucrose affect polymerase chain reaction product formation? Biotechnol Lett 23, 175–178 (2001).

41. Li, Z.H. et al. A chemical-enhanced system for CRISPR-Based nucleic acid detection. Biosens Bioelectron 192 (2021).

42. Spiess, A.N., Mueller, N. & Ivell, R. Trehalose is a potent PCR enhancer: Lowering of DNA melting temperature and thermal stabilization of Taq polymerase by the disaccharide trehalose. Clin Chem 50, 1256–1259 (2004).

43. Jensen, M.A., Fukushima, M. & Davis, R.W. DMSO and Betaine Greatly Improve Amplification of GC-Rich Constructs in De Novo Synthesis. Plos One 5 (2010).

44. Jain, I. et al. Defining the seed sequence of the Cas12b CRISPR-Cas effector complex. Rna Biol 16, 413–422 (2019).

45. Gupta, E., Bajpai, M. & Choudhary, A. Hepatitis C virus: Screening, diagnosis, and interpretation of laboratory assays. Asian J Transfus Sci 8, 19–25 (2014).

46. Madhvi, A. et al. A screen for novel hepatitis C virus RdRp inhibitor identifies a broad-spectrum antiviral compound. Sci Rep-Uk 7 (2017).

47. Spearman, C.W., Dusheiko, G.M., Hellard, M. & Sonderup, M. Hepatitis C. Lancet 394, 1451–1466 (2019).

48. Zeuzem, S. et al. Glecaprevir-Pibrentasvir for 8 or 12 Weeks in HCV Genotype 1 or 3 Infection. N Engl J Med 378, 354–369 (2018).

49. Konerman, M.A. & Lok, A.S. Diagnostic challenges of hepatitis C. JAMA 311, 2536–2537 (2014).

50. Ishizaki, A. et al. Survey of programmatic experiences and challenges in delivery of hepatitis B and C testing in low- and middle-income countries. BMC Infect Dis 17, 696 (2017).

51. Terrault, N.A. Hepatitis C elimination: challenges with under-diagnosis and under-treatment. F1000Res 8 (2019).

52. Hongjaisee, S. et al. Rapid visual detection of hepatitis C virus using a reverse transcription loop-mediated isothermal amplification assay. Int J Infect Dis 102, 440–445 (2021).

53. Mirdita, M. et al. ColabFold: making protein folding accessible to all. Nat Methods 19, 679- + (2022).

54. Bienert, S. et al. The SWISS-MODEL Repository-new features and functionality. Nucleic Acids Research 45, D313–D319 (2017).

55. Zauli, D.A., Menezes, C.L., Oliveira, C.L., Mateo, E.C. & Ferreira, A.C. In-house quantitative real-time PCR for the diagnosis of hepatitis B virus and hepatitis C virus infections. Braz J Microbiol 47, 987–992 (2016).

