## Supplementary Information for "Engineering Highly Thermostable Cas12b via De Novo Structural Analyses for One-Pot Detection of Nucleic Acids"

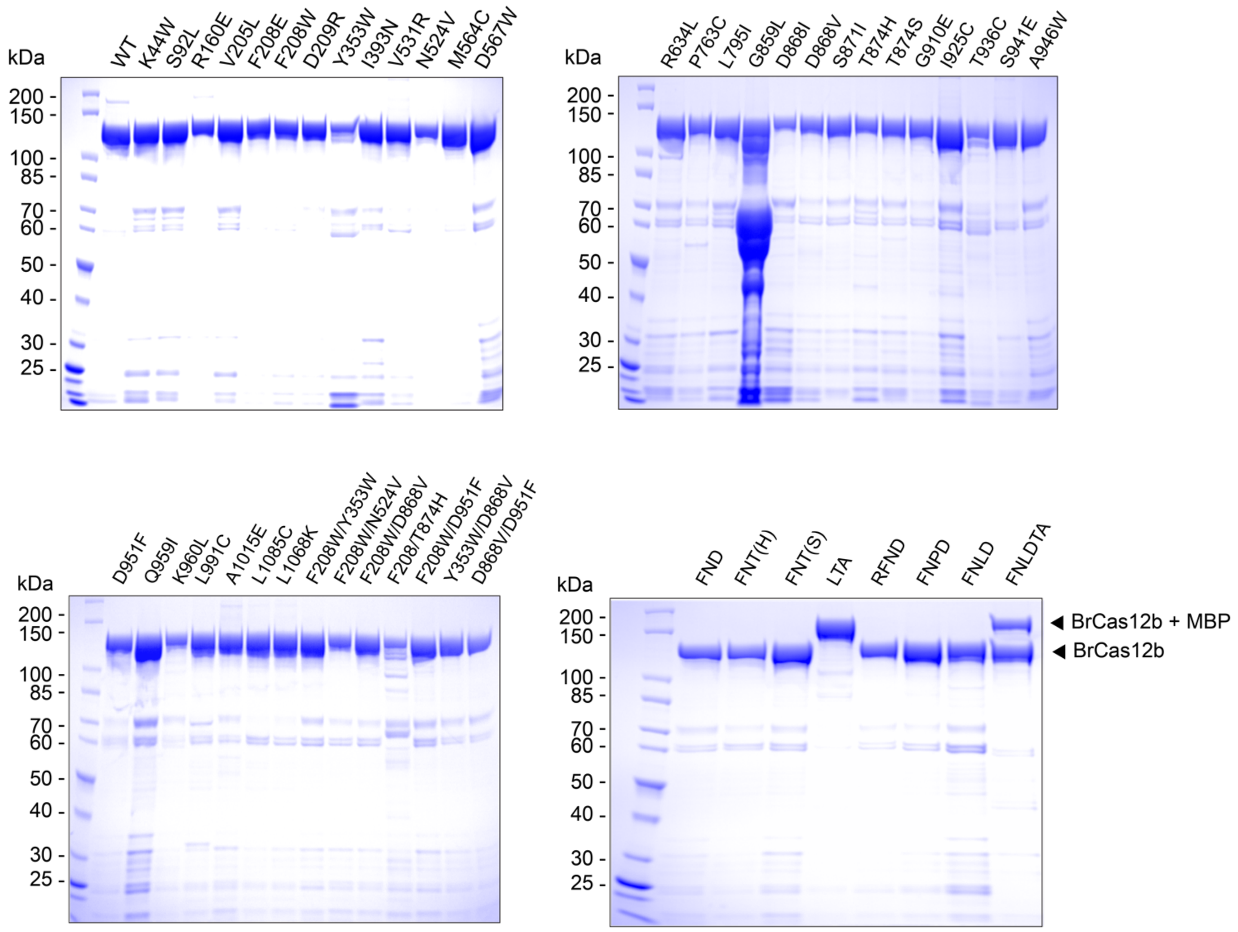


**Figure S1**. SDS-PAGE characterization of all purified BrCas12b variants used in this study. The effectors were either purified from a pET28a$^{+}$ backbone or 6xHis-MBP backbone with a maltose binding protein fused to them.

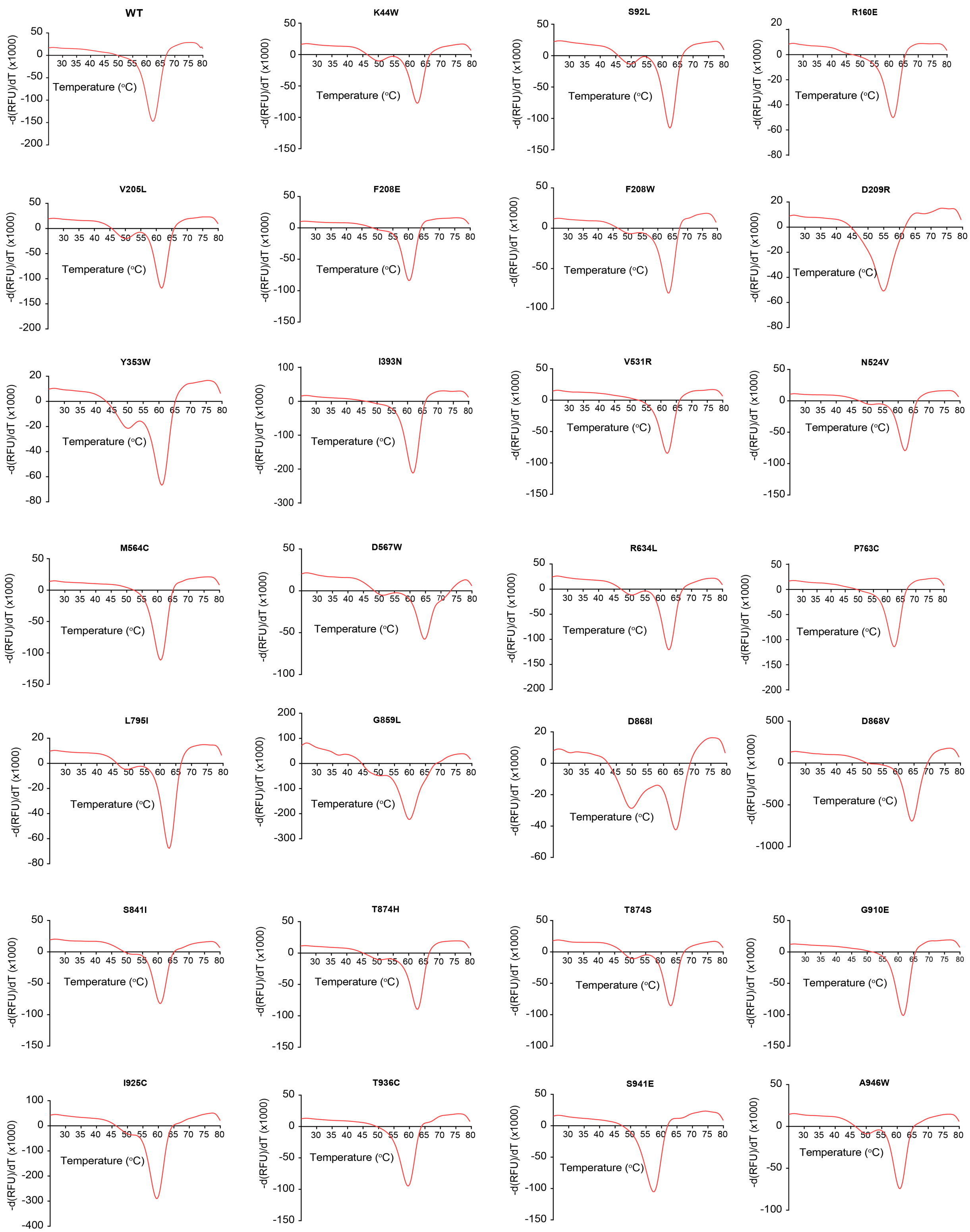
WT
K44W
S92L
R160E
V205L
F208E
F208W
D209R
Y353W
I393N
V531R
N524V
M564C
D567W
R634L
P763C
L795I
G859L
D868I
D868V
S841I
T874H
T874S
G910E
I925C
T936C
S941E
A946W
-d(RFU)/dT (x1000)
Temperature (°C)

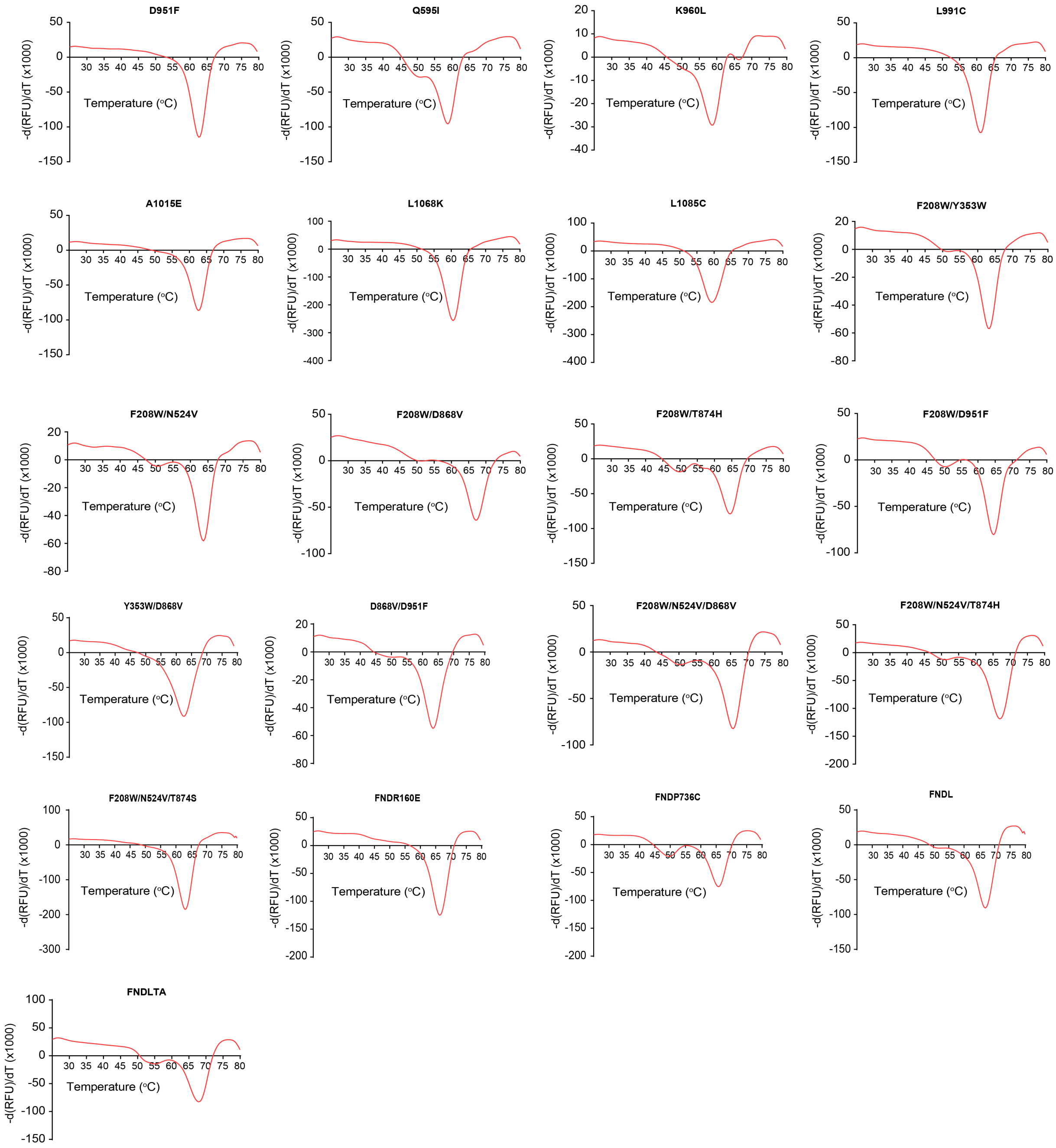


**Figure S2**. Differential scanning fluorometric measurements of all BrCas12b variants used in the study. The curve at each temperature point represents the average of fluorescence over 4 replicates (2 technical replicates per experiment over two experiments). The melting point ($T_m$) was determined as a global minimum of the derivative curve.

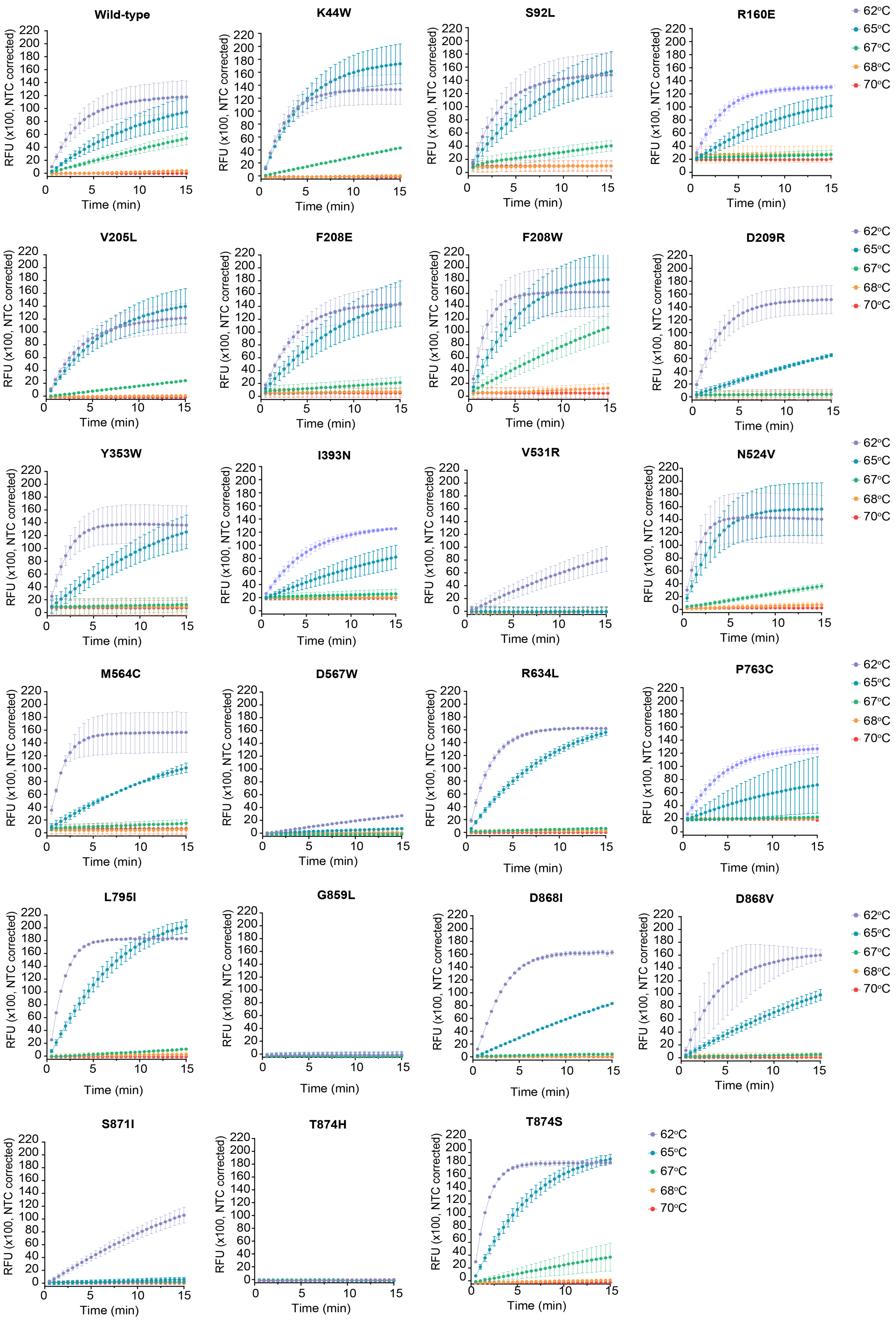

Wild-type
K44W
S92L
R160E
V205L
F208E
F208W
D209R
Y353W
I393N
V531R
N524V
M564C
D567W
R634L
P763C
L795I
G859L
D868I
D868V
S871I
T874H
T874S
62°C
65°C
67°C
68°C
70°C
RFU (x100, NTC corrected)
Time (min)

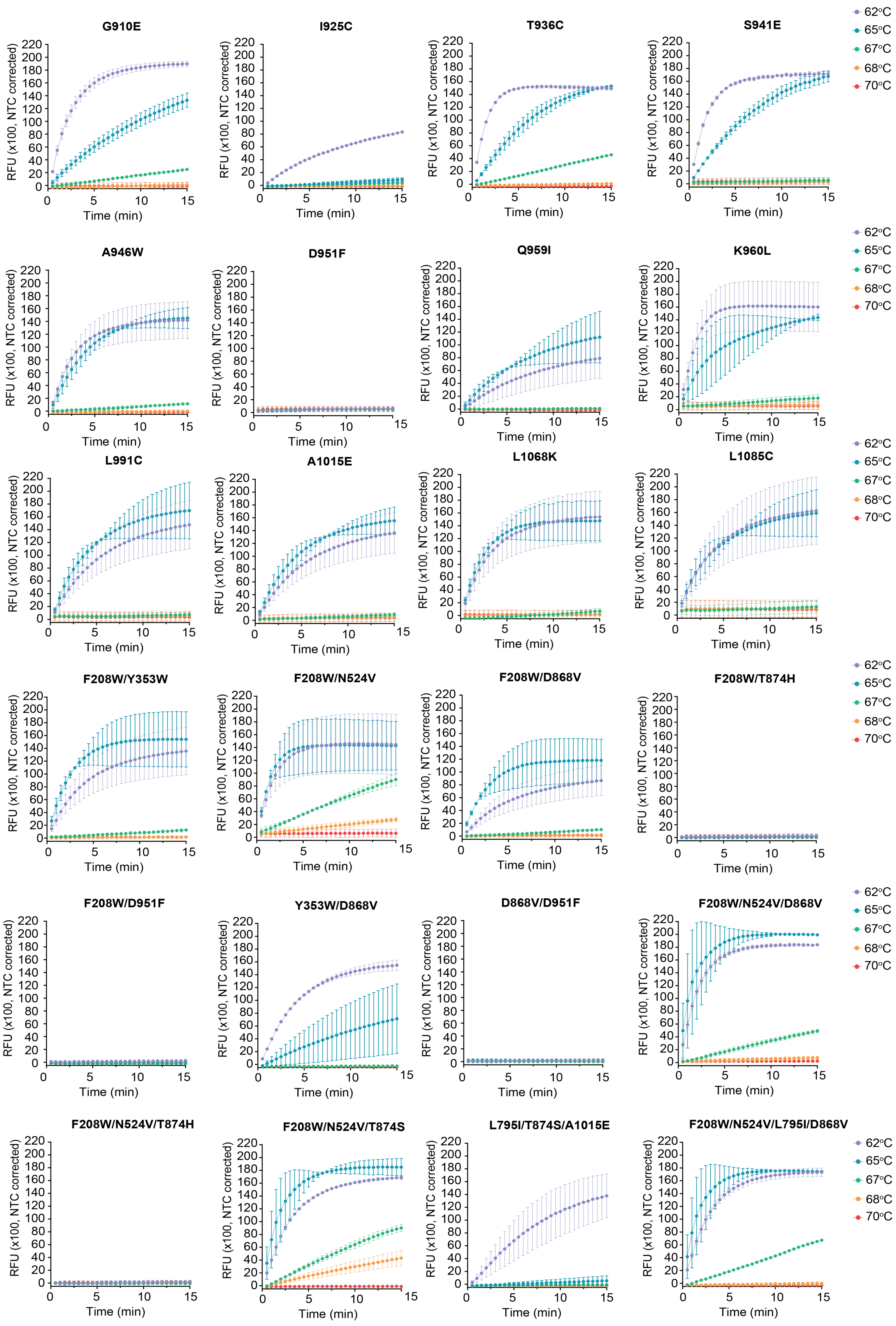

G910E
I925C
T936C
S941E
A946W
D951F
Q959I
K960L
L991C
A1015E
L1068K
L1085C
F208W/Y353W
F208W/N524V
F208W/D868V
F208W/T874H
F208W/D951F
Y353W/D868V
D868V/D951F
F208W/N524V/D868V
F208W/N524V/T874H
F208W/N524V/T874S
L795I/T874S/A1015E
F208W/N524V/L795I/D868V
RFU (x100, NTC corrected)
Time (min)
62°C
65°C
67°C
68°C
70°C

**Figure S3.** Time- and temperature-dependent trans-cleavage activity of BrCas12b variants used in the study. The ribonucleoprotein complex and trans-cleavage reaction were incubated at 62°C, 65°C, 67°C, 68°C, and 70°C, respectively. Error bars represent mean ± s.d., where n = 2 biological replicates.

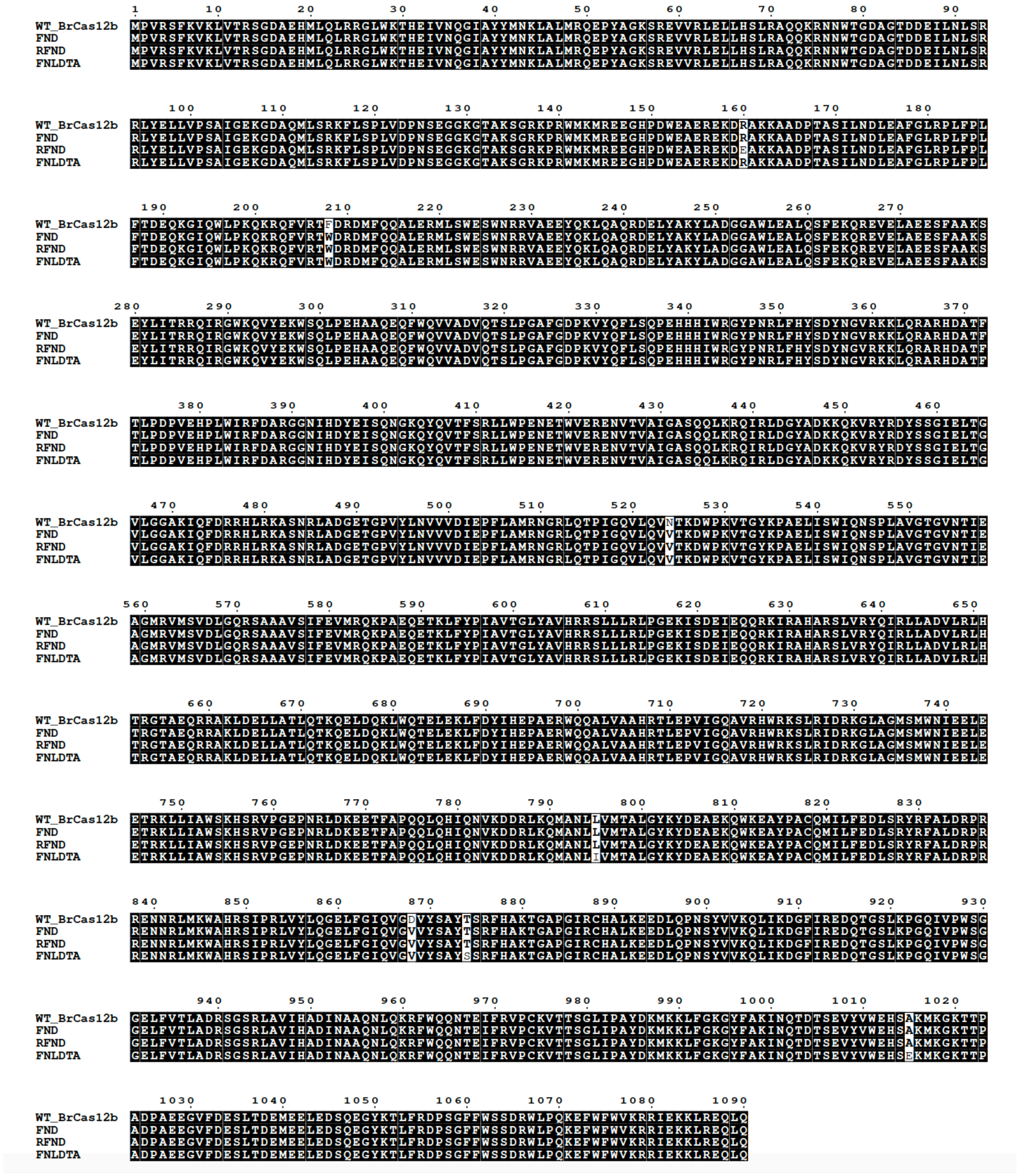

**Figure S4**. Sequence alignment of BrCas12a against promising variants with improved thermostability. FND, RFND, and FNLDTA represent F208W/N524V/D868V, R160E/F208W/N524V/D868V, and F208W/N524V/L795I/D868V/T874S/A1015E, respectively. Protein sequences were aligned with Clustal Omega[1] and input into ESPript 3.0 for visualization[2].

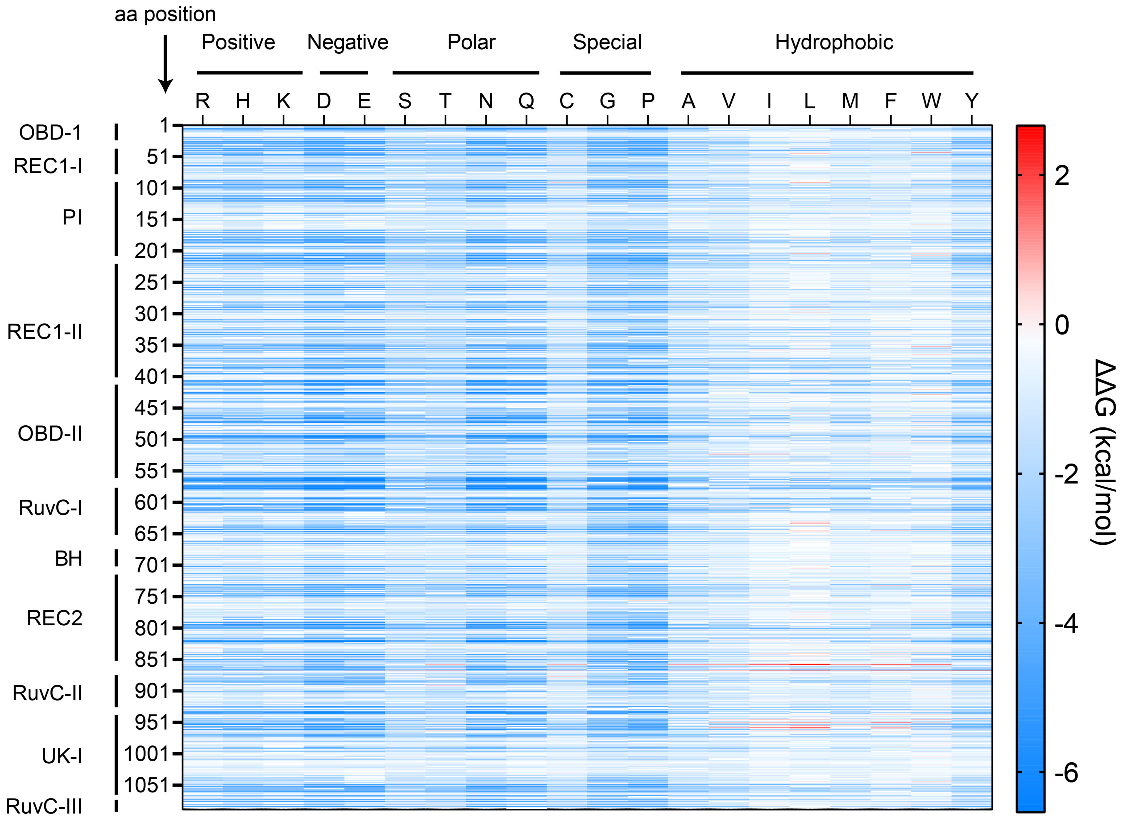


**Figure S5**. Fitness landscape of BrCas12b SWISS-MODEL model calculated by DeepDDG. Each tile represents a computationally predicted change in free energy (ΔΔG) relative to the wild-type BrCas12b for the 20 amino acids.

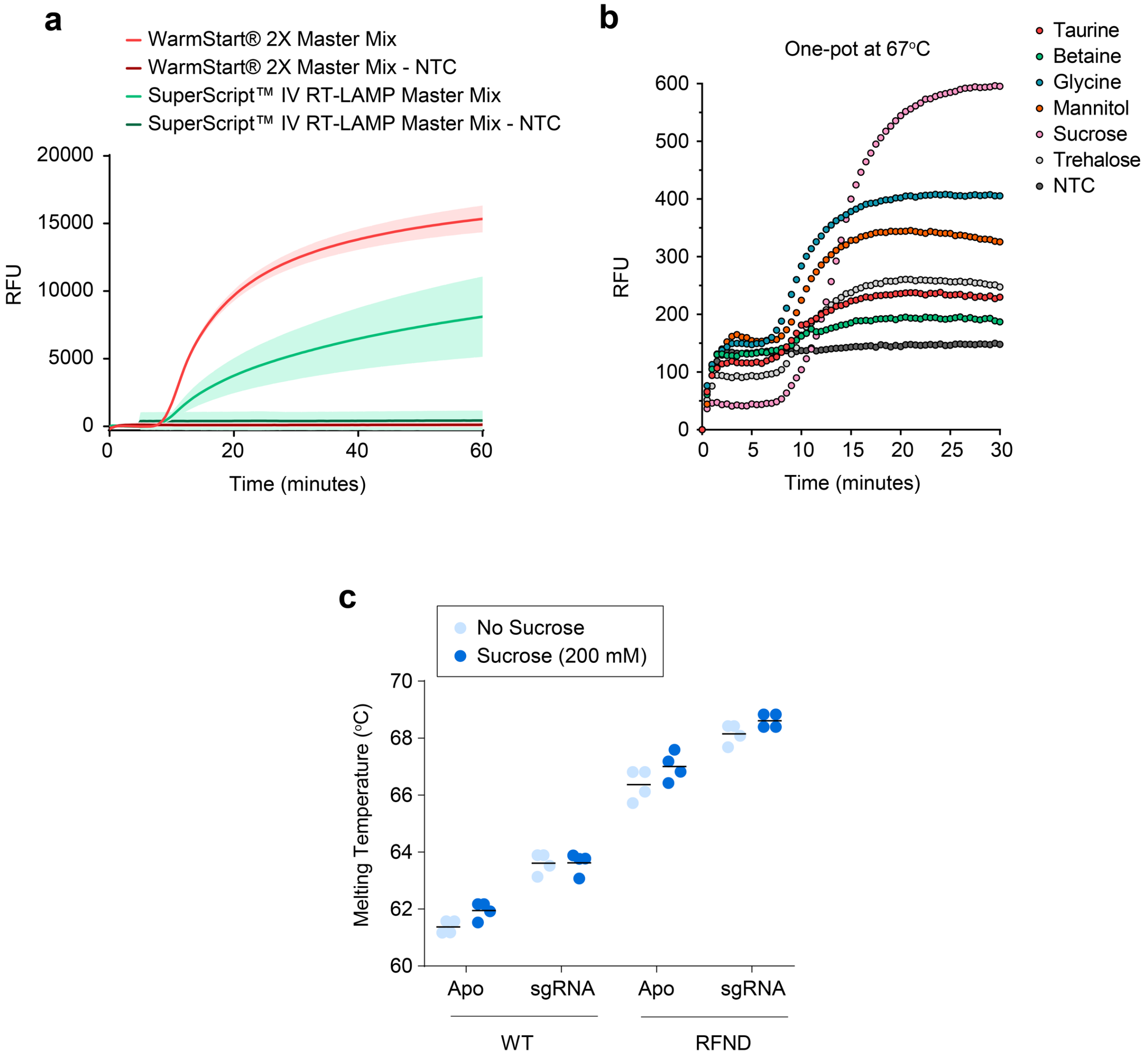


**Figure S6**. BrCas12b detection optimization. (**a**) Compatibility of BrCas12b-based detection reaction with commercially available RT-LAMP master mixes. (**b**) Additive optimization of the one-pot reaction with additives for the FNDLTA variant. Additives were mixed to a final concentration of 50 mM, and the reaction was incubated isothermally at 67°C. Each data point represents an average of triplicates (n = 3 biological replicates).
(**c**) Effects of sucrose on the thermostability of wild-type BrCas12b and the thermally improved RFND variant. Melting temperatures were determined from differential scanning fluorimetry (n = 2 technical replicates per experiment over two experiments). The melting point ($T_m$) was identified as a global minimum of the derivative RFU curve with respect to temperature.

**Table S1. Sequences used in the study**

**RT-LAMP primers**

| Target | Name | Sequence |
|---|---|---|
| N gene of SARS-CoV-2 | N_gene_F3 | AACACAAGCTTTCGGCAG |
| | N_gene_B3 | GAAATTTGGATCTTTGTCATCC |
| | N_gene_FIP | TGCGGCCAATGTTTGTAATCAGCCAA GGAA ATTTTGGGGAC |
| | N_gene_BIP | CGCATTGGCATGGAAGTCACTTTGAT GGC ACCTGTGTAG |
| | N_gene_LF | TTCCTTGTCTGATTAGTTC |
| | N_gene_LB | ACCTTCGGGAACGTGGTT |
| 5' UTR of Hepatitis C[3] | UTR_F3 | CGGGAGAGCCATAGTGGT |
| | UTR_B3 | WGGAWGTGTGCTCATGATGCACG |
| | UTR_FIP | AAATCTCCAGGCATTGAGCGTTTTTGCGGAACCGGTGAGTAC |
| | UTR_BIP | CCGCRAGACYGCTAGCCGAGTTTTACCCTATCAGGCAGTACCAC |
| | UTR_LF | TCGTCCYGGCRATTCCGG |
| | UTR_LB | TAGTGTTGGGTCGCGAAAG |

**Single-guide RNAs**

| Target | Name | Sequence |
|---|---|---|
| N gene of SARS-CoV-2 | Br_sgN_CoV2 | GAAGGUGGUUAGCUACAGGCUGACCAGUGCAGUUGUGUCAUGUGCUACGGUGACCUAACACGUCACUCAGUCACAACGGCUAUCUAUAUUUCCACUAACCAAAGUUAGUGGAAAUGUAGAUGGUUAGCAC**CGAAGAACGCUGAAGCGCUG** |
| 5' UTR of Hepatitis C | Br_sgUTR_HCV | GAAGGUGGUUAGCUACAGGCUGACCAGUGCAGUUGUGUCAUGUGCUACGGUGACCUAACACGUCACUCAGUCACAACGGCUAUCUAUAUUUCCACUAACCAAAGUUAGUGGAAAUGUAGAUGGUUAGCAC**UCCAAGAAAGGACCCGGUCG** |

**RT-qPCR primers[4]**

| Target | Name | Sequence |
|---|---|---|
| 5' UTR of Hepatitis C | HCV_UTR_FOR | AGCGTCTAGCCATGGCGTT |
| | HCV_UTR_REV | GCAAGCACCCTATCAGGCAGT |
| | HCV_UTR_Probe | /56-FAM/TCTGCGGAA/ZEN/CCGGTGAGT/3IABkFQ/ |

Reporters

| Target | Name | Sequence |
|---|---|---|
| Universal | Reporter 1 | /5HEX/TTTTTTTT/3IABkFQ/ |
| | Reporter 2 | /5FAM/TTTTTTTTT/3IABkFQ/ |

**Protein sequences**

| Name | Sequence |
|---|---|

| | |
|---|---|
| AapCas12b | MAVKSIKVKLRLDDMPEIRAGLWKLHKEVNAGVRYYTEWLSLLRQENLYR<br>RSPNGDGEQECDKTAEECKAELLERLRARQVENGHRGPAGSDDELLQLA<br>RQLYELLVPQAIGAKGDAQQIARKFLSPLADKDAVGGLGIAKAGNKPRWV<br>RMREAGEPGWEEEKEKAETRKSADRTADVLRALADFGLKPLMRVYTDSE<br>MSSVEWKPLRKGQAVRTWDRDMFQQAIERMMSWESWNQRVGQEYAKL<br>VEQKNRFEQKNFVGQEHLVHLVNQLQQDMKEASPGLESKEQTAHYVTGR<br>ALRGSDKVFEKWGKLAPDAPFDLYDAEIKNVQRRNTRRFGSHDLFAKLAE<br>PEYQALWREDASFLTRYAVYNSILRKLNHAKMFATFTLPDATAHPIWTRFD<br>KLGGNLHQYTFLFNEFGERRHAIRFHKLLKVENGVAREVDDVTVPISMSE<br>QLDNLLPRDPNEPIALYFRDYGAEQHFTGEFGGAKIQCRRDQLAHMHRRR<br>GARDVYLNVSVRVQSQSEARGERRPPYAAVFRLVGDNHRAFVHFDKLSD<br>YLAEHPDDGKLGSEGLLSGLRVMSVDLGLRTSASISVFRVARKDELKPNS<br>KGRVPFFFPIKGNDNLVAVHERSQLLKLPGETESKDLRAIREERQRTLRQL<br>RTQLAYLRLLVRCGSEDVGRRERSWAKLIEQPVDAANHMTPDWREAFEN<br>ELQKLKSLHGICSDKEWMDAVYESVRRVWRHMGKQVRDWRKDVRSGER<br>PKIRGYAKDVVGGNSIEQIEYLERQYKFLKSWSFFGKVSGQVIRAEKGSRF<br>AITLREHIDHAKEDRLKKLADRIIMEALGYVYALDERGKGKWVAKYPPCQLI<br>LLEELSEYQFNNDRPPSENNQLMQWSHRGVFQELINQAQVHDLLVGTMY<br>AAFSSRFDARTGAPGIRCRRVPARCTQEHNPEPFPWWLNKFVVEHTLDA<br>CPLRADDLIPTGEGEIFVSPFSAEEGDFHQIHADLNAAQNLQQRLWSDFDI<br>SQIRLRCDWGEVDGELVLIPRLTGKRTADSYSNKVFYTNTGVTYYERERG<br>KKRRKVFAQEKLSEEEAELLVEADEAREKSVVLMRDPSGIINRGNWTRQK<br>EFWSMVNQRIEGYLVKQIRSRVPLQDSACENTGDI |
| BrCas12b | MPVRSFKVKLVTRSGDAEHMLQLRRGLWKTHEIVNQGIAYYMNKLALMR<br>QEPYAGKSREVVRLELLHSLRAQQKRNNWTGDAGTDDEILNLSRRLYELL<br>VPSAIGEKGDAQMLSRKFLSPLVDPNSEGGKGTAKSGRKPRWMKMREE<br>GHPDWEAEREKDRAKKAADPTASILNDLEAFGLRPLFPLFTDEQKGIQWL<br>PKQKRQFVRTFDRDMFQQALERMLSWESWNRRVAEEYQKLQAQRDELY<br>AKYLADGGAWLEALQSFEKQREVELAEESFAAKSEYLITRRQIRGWKQVY<br>EKWSQLPEHAAQEQFWQVVADVQTSLPGAFGDPKVYQFLSQPEHHHIW<br>RGYPNRLFHYSDYNGVRKKLQRARHDATFTLPDPVEHPLWIRFDARGGNI<br>HDYEISQNGKQYQVTFSRLLWPENETWVERENVTVAIGASQQLKRQIRLD<br>GYADKKQKVRYRDYSSGIELTGVLGGAKIQFDRRHLRKASNRLADGETGP<br>VYLNVVVDIEPFLAMRNGRLQTPIGQVLQVNTKDWPKVTGYKPAELISWIQ<br>NSPLAVGTGVNTIEAGMRVMSVDLGQRSAAAVSIFEVMRQKPAEQETKLF<br>YPIAVTGLYAVHRRSLLLRLPGEKISDEIEQQRKIRAHARSLVRYQIRLLADV<br>LRLHTRGTAEQRRAKLDELLATLQTKQELDQKLWQTELEKLFDYIHEPAER<br>WQQALVAAHRTLEPVIGQAVRHWRKSLRIDRKGLAGMSMWNIEELEETR<br>KLLIAWSKHSRVPGEPNRLDKEETFAPQQLQHIQNVKDDRLKQMANLLVM<br>TALGYKYDEAEKQWKEAYPACQMILFEDLSRYRFALDRPRRENNRLMKW<br>AHRSIPRLVYLQGELFGIQVGDVYSAYTSRFHAKTGAPGIRCHALKEEDLQ<br>PNSYVVKQLIKDGFIREDQTGSLKPGQIVPWSGGELFVTLADRSGSRLAVI<br>HADINAAQNLQKRFWQQNTEIFRVPCKVTTSGLIPAYDKMKKLFGKGYFA<br>KINQTDTSEVYVWEHSAKMKGKTTPADPAEEGVFDESLTDEMEELEDSQ<br>EGYKTLFRDPSGFFWSSDRWLPQKEFWFWVKRRIEKKLREQLQ |

### References

1. McWilliam, H. et al. Analysis Tool Web Services from the EMBL-EBI. *Nucleic Acids Res* **41**, W597-600 (2013).
2. Robert, X. & Gouet, P. Deciphering key features in protein structures with the new ENDscript server. *Nucleic Acids Res* **42**, W320-324 (2014).
3. Hongjaisee, S. et al. Rapid visual detection of hepatitis C virus using a reverse transcription loop-mediated isothermal ampli fi cation assay. *Int J Infect Dis* **102**, 440-445 (2021).
4. Zauli, D.A., Menezes, C.L., Oliveira, C.L., Mateo, E.C. & Ferreira, A.C. In-house quantitative real-time PCR for the diagnosis of hepatitis B virus and hepatitis C virus infections. *Braz J Microbiol* **47**, 987-992 (2016).
